# Determinants of Subjective Well-Being in Taiwan: A Machine Learning and SHAP Analysis

**DOI:** 10.1101/2025.10.05.25337343

**Authors:** Melody Hsiao-San Yeh, Chun-Tung Kuo, Shu-Hui Hsieh, Tzu-Pin Lu, Vincent Cheng-Sheng Li, John Tayu Lee

**Affiliations:** Institute of Health Policy and Management, College of Public Health, National Taiwan University, Taipei, Taiwan (R.O.C.); Research Center for Humanities and Social Sciences, Academia Sinica, Taipei, Taiwan (R.O.C.); Institute of Health Behaviors and Community Sciences, College of Public Health, National Taiwan University, Taipei, Taiwan (R.O.C.); Institute of Health Data Analytics and Statistics, College of Public Health, National Taiwan University, Taipei, Taiwan (R.O.C.)

**Keywords:** Machine Learning, Wellbeing, Explainable Machine Learning, Social Determinants

## Abstract

This study aimed to identify and rank key determinants of subjective well-being (SWB), examine threshold values, distinguish protective from motivational roles, assess non-linear associations, and evaluate the performance of machine learning models in predicting SWB. We conducted a cross-sectional machine learning study using data from 10,712 participants in Taiwan, derived from a nationally representative 2024 survey. SHAP analysis was applied to interpret model outputs. We identified five key determinants: family, interpersonal relationships, health, life goal clarity, and financial stability. Family relationship satisfaction, interpersonal relationship satisfaction, and health are protective factors, all of which show nonlinear associations with SWB, including pronounced threshold effects: when scores exceed 5–6 on a 10-point scale, SWB increases sharply, underscoring the importance of reaching sufficient intervention intensity. In contrast, financial safety and goal clarity function as motivational factors. Gradient Boosting demonstrated the strongest predictive performance. These findings highlight several actionable levers for enhancing SWB, including strengthening social relationships, improving health, clarifying life goals, and ensuring financial safety. Importantly, the presence of threshold effects suggests that benefits are not uniformly distributed but disproportionately realized among individuals with lower baseline conditions—often the most vulnerable.

## INTRODUCTION

Subject well-being (SWB) is a universal cornerstone of human flourishing, not a privilege reserved for affluent societies (Ryff, 2024). It anchors modern public governance, driving social equity, resilience, and sustainable progress in an era of unprecedented complexity (Fioramonti et al., 2022). Against the backdrop of global population aging, escalating mental health challenges, and widen health disparities, SWB has ascended to a pivotal role in public policy (Compton, 2023; Whyle & Olivier, 2020). The global institutions such as the OECD and the United Nations have explicitly stated well-being at the core of governance frameworks, with Sustainable Development Goals (SDGs) 3 “good health and wellbeing” underscoring its critical role in fostering equitable and resilient and equitable development (OECD, 2013; United Nations, 2024).

SWB is conceptualized as a multidimensional construct, shaped by a dynamic interplay of factors. These include psychological resilience and social connectedness (Azizan & Mahmud, 2018; Diener et al., 1999; Dury et al., 2021), access to material and educational resources (Azizan & Mahmud, 2018; Biswas-Diener & Diener, 2001; Lamu & Olsen, 2016; OECD, 2013; Uwannah et al., 2021), family dynamics and relationship intimacy (Diener et al., 2018; OECD, 2013), and neighborhood attributes such as accessibility and perceived safety (Han et al., 2024; OECD, 2023; Wang et al., 2019). Health-related factors- including physical condition, multimorbidity, and engagement in health-promoting behaviors- have also been identified as key contributors to wellbeing (Diener et al., 1999; OECD, 2013).

Machine learning has significantly advanced the prediction of subjective well-being (SWB) by uncovering complex determinant patterns. For instance, Panicheva et al. used machine learning and deep learning methods utilizing mental health apps data to predict wellbeing (Panicheva et al., 2022). Zhang et al., González-Carrasco et al., and King et al. utilized gradient boosting approaches to predict adolescent SWB with high accuracy(González-Carrasco et al., 2024; King et al., 2024; Zhang et al., 2019). More recently, Oparina et al. (2025) scaled tree-based models to over one million respondents, demonstrating their superiority over linear methods in capturing non-linear relationships(Oparina et al., 2025). Nevertheless, important limitations remain. Most studies are grounded in Western contexts, limiting their cultural applicability.

This study is grounded in complementary theoretical frameworks, including Keyes’ model of psychological, emotional, and social well-being, Maslow’s hierarchy of needs, and the Theory of Fundamental Causes, which posits that well-being depends on foundational resources (e.g., health and financial security) as well as higher-order capacities such as purpose and social integration (Maslow, 1943; Robitschek & Keyes, 2009). These frameworks inform our classification of SWB determinants as either protective or motivational and guide our exploration of non-linear and threshold effects.

Simultaneously, this study adopts the Social Ecological Model (SEM) to structure the analysis (Center for Leadership Education in Maternal and Child Public Health, 2015). The model views well-being not as an isolated phenomenon, but as shaped by interactions across nested social layers (Center for Leadership Education in Maternal and Child Public Health, 2015). SEM delineates five levels: individual, interpersonal, organizational, community, and societal (Center for Leadership Education in Maternal and Child Public Health, 2015).

By integrating machine learning with established theory, we aim to combine predictive performance with social explanation. Taiwan’s sociocultural context, characterized by strong filial norms, multigenerational living, and religious adherence, shape relational expectations and support structures in ways that may intensify the influence of family satisfaction and interpersonal trust (OLIVEIRA et al., 2019; Villani et al., 2019). Additionally, rapid urbanization, regional inequality, and evolving civic norms contribute to variation in perceived safety and participation, underscoring the need for culturally situated analysis (Cacciatore et al., 2025; Dang et al., 2022).

To date, no study in Taiwan has applied explainable machine learning methods—such as SHAP (SHapley Additive exPlanations)—to assess SWB. This approach enables quantification of determinant contribution, directionality, and threshold effects. Our study pursues four objectives: (1) to evaluate performance of a series of machine learning models for SWB prediction; (2) to identify and rank key determinants and their relative contributions; (3) to examine threshold values and distinguish protective from motivational factors; and (4) to assess potential non-linear relationships between determinants and SWB.

## METHODS

### Data and Samples

The data used data from the 2024 national well-being survey conducted by the Center for Survey Research (CSR) at the Research Center for Humanities and Social Sciences, Academia Sinica in Taiwan. Data collection took place between October 11 and November 29, 2024. Respondents were adults aged 18 years and older residing in Taiwan. The survey employed a two-stage recruitment strategy to maximize representativeness. In the first stage, invitations were sent via email to members of CSR’s existing panel who had previously consented to participate in follow-up surveys. In the second stage, a professional fieldwork agency distributed the questionnaire link via Short Message Service (SMS) to additional individuals outside the panel, broadening demographic diversity. Of the 12,260 individuals who completed the survey, 10,712 provided complete responses and were included in the final analytic sample (Chang, 2023).

### Measurements

The primary outcome variable in this study is the SWB category. SWB was operationalized as the mean of two core components: self-reported happiness and life satisfaction, each measured on an 11- point scale ranging from 0 to 10. This composite index reflects individuals’ cognitive and affective evaluation of their overall life experience (Nisbet et al., 2011; Somarriba Arechavala et al., 2022).

Respondents were classified into two groups based on the distribution of SWB scores: (1) a lower SWB group (scores 0–6, inclusive), and (2) a higher SWB group (scores above 6). A cutoff at 6 achieved optimal balance between data distribution and conceptual interpretability, helping to reduce bias. This binary categorization enhances interpretability while retaining empirical sensitivity to population-level variation in well-being.

Regarding the independent variables, this study is grounded in the Social Ecological Model, which organizes influences on well-being across five levels. At the individual level, the relevant dimensions are health and satisfaction, along with psychological and behavioral attributes. At the interpersonal level, the key dimension is social participation. At the organizational level, the dimension includes indicators of socioeconomic status. At the community level, the dimension captures aspects of the neighborhood environment. At the societal level, the associated dimension encompasses demographic characteristics. A comprehensive list and definition of these variables is provided in *Appendix Table A.1*.

### Preprocessing of Independent Variables

A systematic preprocessing strategy was implemented to ensure data integrity and analytical consistency. The overall workflow is depicted in *Fig. 1* Categorical variables were encoded using one-hot (nominal) or ordinal (ordered) schemes to minimize multicollinearity. Continuous variables were standardized to ensure comparability across features. Missing values were addressed through casewise deletion to maintain the validity of the final dataset. Feature engineering steps included the removal of extreme outliers and the exclusion of variables with disproportionate influence on model behavior, thereby reducing the risk of overfitting. Derived variables were also created to enhance predictive utility—for example, body mass index (BMI) was calculated from reported height and weight, and then categorized based on established clinical cutoffs. Selected continuous variables were additionally recoded into categorical formats to improve model interpretability and alignment with policy-relevant thresholds.

**Fig. 1.**
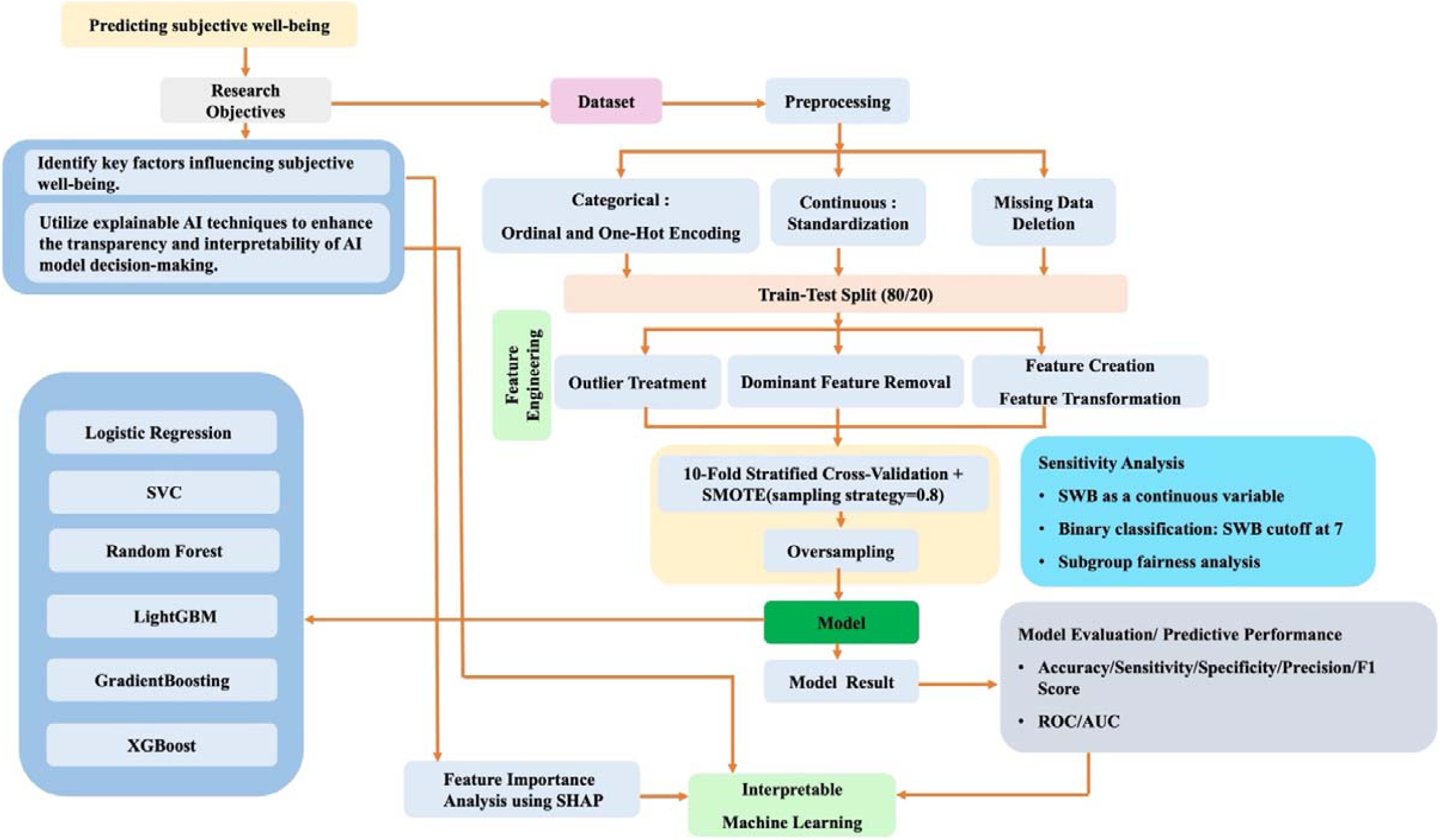
Workflow Diagram for Machine Learning Research Analysis. This figure illustrates the full analytic process, including data preprocessing, machine learning model training, performance evaluation, and SHAP-based model interpretation.

### Machine Learning Algorithms

A supervised classification task was conducted to predict SWB categories. The dataset was split into training and testing sets using an 80/20 stratified sampling approach to maintain class distribution. A 10-fold stratified cross-validation was performed on the training data to assess model robustness.

During each fold of the cross-validation process, Synthetic Minority Over-sampling Technique (SMOTE) was applied to the training subset to address the class imbalance issue, thereby reducing potential bias caused by disproportionate category distributions (Khan et al., 2024). Five machine learning models were implemented: Logistic Regression, Random Forest, LightGBM, Gradient Boosting, and XGBoost (Bozorgmehr & Weltermann, 2023; Khan et al., 2024; Tseng et al., 2012). Model hyperparameters were initially set according to empirical experience to enhance robustness and reduce the risk of overfitting. For example, in the Random Forest model, the number of decision trees was set to 100, and the maximum tree depth was limited to 10. For the ensemble algorithms (LightGBM, Gradient Boosting, and XGBoost), key parameters—including learning rate, tree depth, and regularization terms—were tuned based on existing literature and practical experience to enhance both performance and generalizability.

### Model Evaluation and Predictive Performance

Model performance was assessed using a suite of standard classification metrics, each capturing a distinct aspect of predictive validity. Accuracy measured the overall proportion of correct classifications. Sensitivity (or recall) quantified the model’s ability to correctly identify individuals with higher SWB, while specificity assessed its capacity to recognize those with lower SWB. Precision reflected the proportion of true positives among predicted positives, and the F1-score, as the harmonic mean of precision and recall, offered a balanced metric particularly suited to imbalanced class distributions. To further assess discriminatory power, the receiver operating characteristic (ROC) curve and the area under the curve (AUROC) were employed (Bozorgmehr & Weltermann, 2023). AUROC values closer to 1.0 indicate superior ability to distinguish between outcome classes.

### Feature Importance Analysis

To enhance model interpretability, Shapley Additive Explanations (SHAP) were employed to quantify the contribution of individual features to the model’s predictions. SHAP values were computed using the TreeExplainer algorithm within the LightGBM framework, with categorical variables passed using the native category data type to preserve structural integrity. A series of SHAP visualizations were generated to facilitate interpretation. These included global feature importance plots, summary plots, and dependence plots for the top five predictors.

### Sensitivity Analysis

To assess the robustness of the findings, three complementary sensitivity analyses were conducted. First, SWB was modeled as a continuous outcome to evaluate the consistency of predictor effects under an alternative functional specification. Second, a revised binary classification was applied using a more conservative threshold, reclassifying respondents with SWB scores above seven as the higher well-being group. Third, subgroup fairness analysis was undertaken to examine equity in model performance across key sociodemographic strata. This analysis employed three fairness metrics—equalized odds difference, disparate impact, and equal opportunity difference—to evaluate whether performance disparities were mitigated following model debiasing procedures (Kearns et al., 2019; Shui et al., 2022).

### Software

All analyses were conducted using Python (version 3.12.2) and Stata (version 18).

### Ethics

This study was approved by the Institutional Review Board for Humanities & Social Science Research at Academia Sinica (AS-IRB-HS 02-19025(R15)) and the Research Ethics Committee of National Taiwan University (No. 202411HS043). The survey data were retrospectively de-identified to ensure compliance with ethical research guidelines. For the questionnaire data used in this study, all respondents signed the informed consent form. The data analysis was conducted after de-identification, in order to safeguard the privacy of the respondents and uphold data confidentiality principles.

## RESULTS

### Descriptive Summary

The sample comprised 10,712 respondents, with 78.57% reporting current employment. In terms of monthly income, 20.86% earned less than NTD 30,000 (approximately USD 960), 34.18% earned between NTD 30,000 and NTD 50,000 (USD 960–1,600), and 39.80% earned above NTD 50,000 (USD ≥1,600). Regarding educational attainment, 48.17% of respondents held a university degree, and an additional 23.20% had completed postgraduate education (master’s or doctoral level) *(see Tables 1a and 1b)*.

**Table 1a.** Summary Statistics of Continuous Variables.

| **N=10,712** | **Mean±SD** | **Min** | **Max** | **Median** | **IQR** |
| --- | --- | --- | --- | --- | --- |
| **Demographic Variables** |  |  |  |  |  |
| **Age** | 42.95±12.54 | 18 | 70 | 43 | 19 |
| **Health and Satisfaction Variables** |  |  |  |  |  |
| **Health Status** | 6.39±1.92 | 0 | 10 | 7 | 3 |
| **Mental Health** | 6.62±2.18 | 0 | 10 | 7 | 3 |
| **Job Satisfaction** | 6.03±2.33 | 0 | 10 | 6 | 3 |
| **Family Satisfaction** | 6.96±2.18 | 0 | 10 | 7 | 2 |
| **Interpersonal Relationship Satisfaction** | 6.80±1.98 | 0 | 10 | 7 | 2 |
| **Financial Satisfaction** | 5.70±2.41 | 0 | 10 | 6 | 3 |
| **Family Relationship Satisfaction** | 6.93±2.15 | 0 | 10 | 7 | 2 |
| **Psychological and Behavioral Variables** |  |  |  |  |  |
| **Public Welfare Commitment** | 5.74±2.40 | 0 | 10 | 6 | 2 |
| **Delayed Gratification** | 6.12±2.22 | 0 | 10 | 6 | 3 |
| **Financial Safety** | 5.46±2.90 | 0 | 10 | 5 | 5 |
| **Personal Safety** | 5.91±2.68 | 0 | 10 | 6 | 4 |
| **Life Purpose** | 6.50±2.03 | 0 | 10 | 7 | 3 |
| **Life Goal** | 6.21±2.41 | 0 | 10 | 6 | 3 |
| **Home Return Days** | 5.73±1.93 | 0 | 7 | 7 | 2 |

**Table 1b.** Summary Statistics of Categorical Variables.

| **N=10,712** | **Frequency(n)** | **Percent(%)** |
| --- | --- | --- |
| **Outcome Variable** |  |  |
| **SWB Category** |  |  |
| Lower SWB Group | 4621 | 43.14 |
| Higher SWB Group | 6091 | 56.86 |
| **Demographic Variables** |  |  |
| **Gender** |  |  |
| Male | 4955 | 46.26 |
| Female | 5757 | 53.74 |
| **Geographical Regions** |  |  |
| Northern Region | 6049 | 56.47 |
| Central Region | 1963 | 18.33 |
| Southern Region | 2486 | 23.21 |
| Eastern Region | 214 | 2 |
| **Religion** |  |  |
| No Religion | 2642 | 24.66 |
| Folk Religion | 4896 | 45.71 |
| Buddhism/Taoism | 2202 | 20.56 |
| I-Kuan Tao/ Catholicism/ Christianity/ Islam | 972 | 9.07 |
| **BMI Group** |  |  |
| Underweight | 602 | 5.62 |
| Normal | 5200 | 48.54 |
| Overweight | 2578 | 24.07 |
| Obese | 2332 | 21.77 |
| **Marital Status** |  |  |
| Unmarried | 5424 | 50.63 |
| Married | 5288 | 49.37 |
| **Socioeconomic Variables** |  |  |
| **Working State** |  |  |
| Not Working | 2296 | 21.43 |
| Currently Working | 8416 | 78.57 |
| **Income** |  |  |
| No Income | 553 | 5.16 |
| Less Than NTD 30,000 | 2235 | 20.86 |
| NTD 30,000-50,000 | 3661 | 34.18 |
| NTD 50,000 or More | 4263 | 39.8 |
| **Educational Attainment** |  |  |
| Under University | 3067 | 28.63 |
| University | 5160 | 48.17 |
| Master/PhD | 2485 | 23.2 |
| **Neighborhood Environment Variables** |  |  |
| **Nearby Shops** |  |  |
| Yes | 10028 | 93.61 |
| No | 684 | 6.39 |
| **Nearby Transit** |  |  |
| Yes | 8665 | 80.89 |
| No | 2047 | 19.11 |
| **Nearby Relaxation Spots** |  |  |
| Yes | 8035 | 75.01 |
| No | 2677 | 24.99 |
| **Traffic Safety** |  |  |
| Safe | 4218 | 39.38 |
| Unsafe | 6494 | 60.62 |
| **Crime Safety** |  |  |
| Safe | 1372 | 12.81 |
| Unsafe | 9340 | 87.19 |
| **Social Participation Variables** |  |  |
| **Charity Participation** |  |  |
| Never | 3364 | 31.4 |
| Rarely | 4553 | 42.5 |
| Sometimes | 2268 | 21.17 |
| Often | 527 | 4.92 |
| **Community Improvement** |  |  |
| Never | 5917 | 55.24 |
| Rarely | 3529 | 32.94 |
| Sometimes | 1127 | 10.52 |
| Often | 139 | 1.3 |
| **Volunteer Work** |  |  |
| Never | 5859 | 54.7 |
| Rarely | 3199 | 29.86 |
| Sometimes | 1135 | 10.6 |
| Often | 519 | 4.85 |
| **Community Meetings** |  |  |
| Never | 5891 | 54.99 |
| Rarely | 3328 | 31.07 |
| Sometimes | 1215 | 11.34 |
| Often | 278 | 2.6 |
| **Social Issue Participation** |  |  |
| Never | 6482 | 60.51 |
| Rarely | 3132 | 29.24 |
| Sometimes | 944 | 8.81 |
| Often | 154 | 1.44 |
| **Political Activity** |  |  |
| Never | 7777 | 72.6 |
| Rarely | 2245 | 20.96 |
| Sometimes | 606 | 5.66 |
| Often | 84 | 0.78 |
| **Religious Donations** |  |  |
| Never | 5355 | 49.99 |
| Rarely | 3103 | 28.97 |
| Sometimes | 1629 | 15.21 |
| Often | 625 | 5.83 |
| **Non Religious Donations** |  |  |
| Never | 4778 | 44.6 |
| Rarely | 3534 | 32.99 |
| Sometimes | 1777 | 16.59 |
| Often | 623 | 5.82 |
| **Helping Others** |  |  |
| Never | 2434 | 22.72 |
| Rarely | 4408 | 41.15 |
| Sometimes | 3034 | 28.32 |
| Often | 836 | 7.8 |
| **Public Opinion Polling** |  |  |
| Strongly Disagree | 483 | 4.51 |
| Disagree | 1402 | 13.09 |
| Neutral | 4145 | 38.69 |
| Agree | 3981 | 37.16 |
| Strongly Agree | 701 | 6.54 |
| **Civic Expression** |  |  |
| Strongly Disagree | 112 | 1.05 |
| Disagree | 403 | 3.76 |
| Neutral | 2793 | 26.07 |
| Agree | 6152 | 57.43 |
| Strongly Agree | 1252 | 11.69 |

Regarding the outcome variable, 43.14% of participants were classified into the lower SWB group, while 56.86% were categorized as having higher SWB. In terms of demographic characteristics, 46.26% of respondents were male. Participants ranged in age from 18 to 70 years, with a mean age of 42.95 years (SD = 12.54) and a median of 43 years. Based on body mass index (BMI), approximately half of the sample fell within the normal weight range, while 24.07% were classified as overweight and 21.77% as obese. A total of 49.37% were married. Regarding religious affiliation, 24.66% identified as having no religion, whereas 45.71% adhered to folk religious traditions. Geographically, 56.47% of respondents resided in the northern region of Taiwan, followed by 23.21% in the south and 18.33% in the central region.

With respect to the neighborhood environment, 93.61% of respondents reported access to shops within a 10-minute walking distance, and 75.01% had nearby relaxation spaces. Nevertheless, 60.62% perceived their traffic surroundings as unsafe. Within the domain of social participation, responses related to charitable activities showed that 42.50% of participants reported rare engagement, 31.40% had never participated, and 21.17% participated occasionally. In community improvement initiatives, 55.24% indicated no participation, and only 10.52% engaged sometimes. Similarly, participation in volunteer work, community meetings, social issue campaigns, and political events remained limited, with over half of respondents reporting no involvement. Donation behavior exhibited slight variation: religious donations were marginally more common than non-religious ones. Nonetheless, civic attitudes show that 43.7% of respondents agreed or strongly agreed on the value of public opinion polling, and approximately 70% affirmed the importance of civic expression in public affairs. In the domain of health and satisfaction, the mean (SD) values were as follows: self-rated health status 6.39 (1.92), mental health 6.62 (2.18), job satisfaction 6.03 (2.33), family satisfaction 6.96 (2.18), interpersonal relationship satisfaction 6.80 (1.98), financial satisfaction 5.70 (2.41), and family relationship satisfaction 6.93 (2.15). Within the psychological and behavioral domain, the mean (SD) scores were as follows: public welfare commitment, 5.74 (2.40); delayed gratification, 6.12 (2.22); financial safety, 5.46 (2.90); personal safety, 5.91 (2.68); life purpose, 6.50 (2.03); and life goals, 6.21 (2.42).

### Model Evaluation and Predictive Performance of Machine Learning Models

*Table 2* presents the classification performance of multiple machine learning models across several evaluation metrics, including accuracy, sensitivity, specificity, precision, F1 score, and AUROC. Model accuracy ranged from 0.817 to 0.824. Gradient Boosting achieved the highest accuracy (0.824). Other models, including ensemble approaches, demonstrated slightly lower classification performance. With respect to sensitivity, Gradient Boosting again performed best (0.858). Specificity values ranged from 0.766 to 0.781 across models. In terms of precision, the XGBoost model outperformed all others, achieving a value of 0.837, followed by LightGBM and Gradient Boosting (both 0.836). The highest F1 score was observed for the Gradient Boosting model (0.847), with XGBoost and Logistic Regression performing comparably at 0.845. To assess model discrimination, ROC curves and AUROC values were examined *(See Table 2 and Fig. 2)*. XGBoost showed the highest AUROC (0.905), followed closely by Gradient Boosting (0.904). All models demonstrated similar ability to distinguish between high- and low-SWB groups across both training and testing sets.

**Fig. 2.**
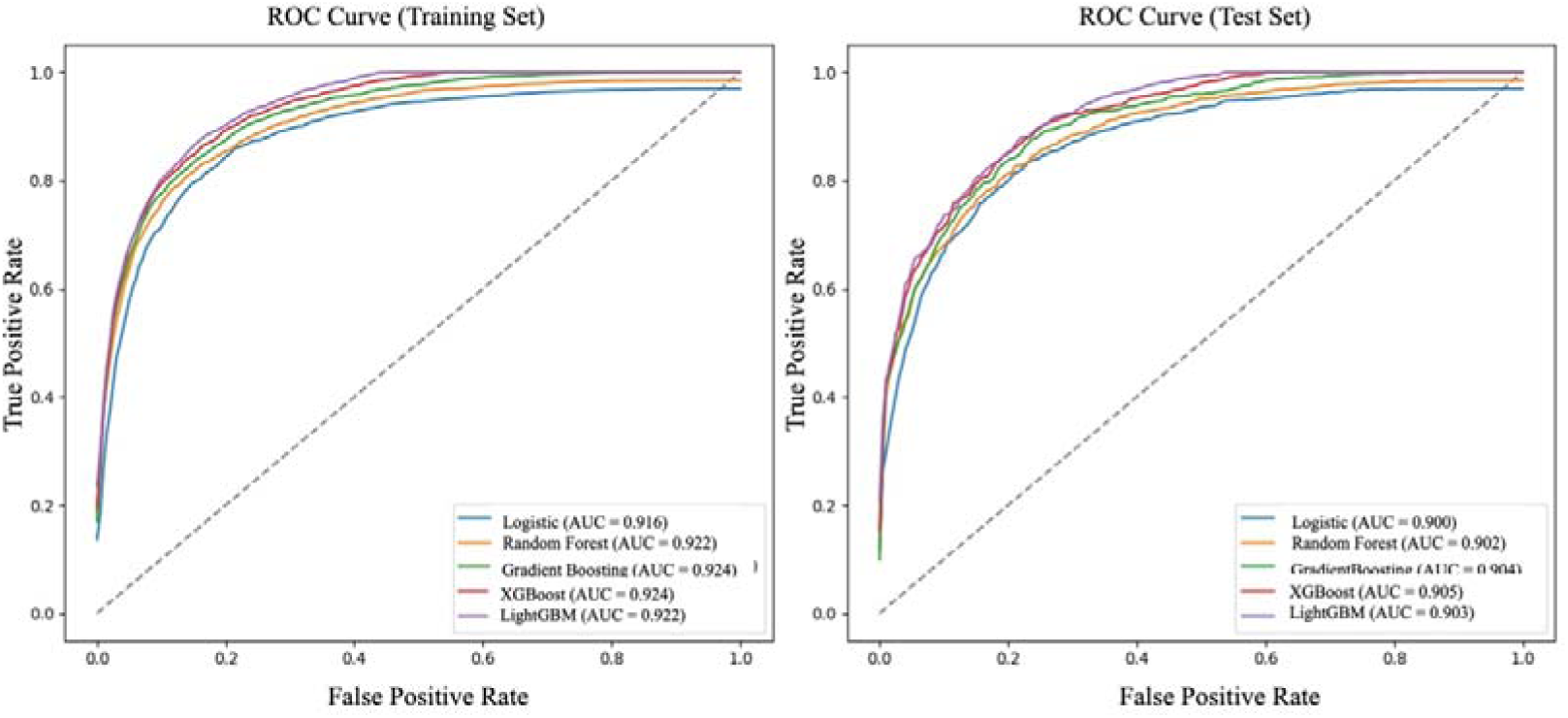
ROC Curves for Training and Testing Sets. The figure presents Receiver Operating Characteristic curves for five machine learning models. The left panel shows ROC curves for the training set, and the right panel for the test set. XGBoost achieved the highest Area Under the Curve values.

**Table 2.**
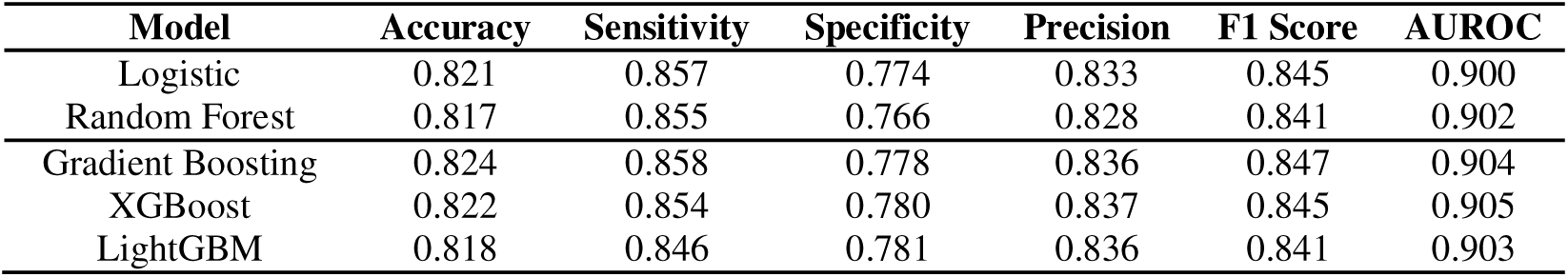
Model Evaluation and Predictive Performance of Machine Learning Models.

### Feature Importance Analysis

*Fig. 3* presents a ranked visualization of the most influential determinants of SWB, based on SHAP values. Variables are color-coded by domain: health and satisfaction (red), psychological and behavioral (brown), socioeconomic (green), demographic (blue), neighborhood environment (orange), and social participation (purple).

**Fig. 3.**
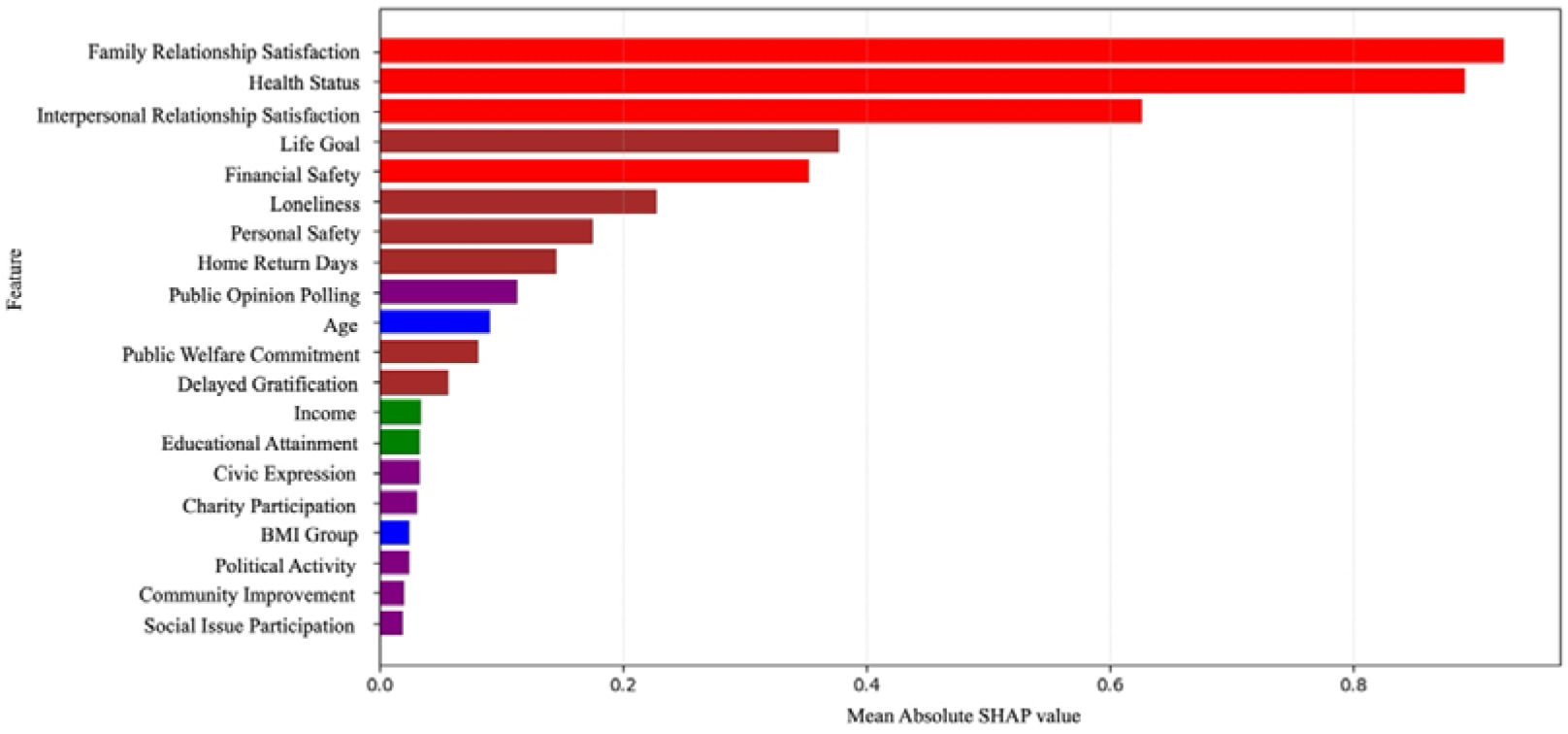
Top Features by Aggregated SHAP Value. The figure shows the top predictors of subjective well-being based on SHAP values from the LightGBM model. Higher SHAP values reflect greater model contribution. Variables are color-coded by domain: health/satisfaction (red), psychological/behavioral (brown), socioeconomic (green), demographic (blue), neighborhood (orange), and social participation (purple).

Among all predictors, family relationship satisfaction was ranked highest. Other top-ranked variables included self-rated health, interpersonal relationship satisfaction, life goal clarity, and financial safety. Variables from the psychological and behavioral domain—such as loneliness, perceived personal safety, and frequency of early return home were also among the top contributors. Overall, features from the health and satisfaction and psychological and behavioral domains accounted for nearly 70% of total feature importance, indicating that internal and interpersonal experiences are the primary contributors of SWB. In contrast, traditionally emphasized socioeconomic and demographic variables—including age, objective income, and educational attainment—showed lower importance scores.

### Direction and Effect of SWB Determinants (From SHAP)

*Fig. 4* (SHAP summary plot) illustrates the directional impact of key predictors on SWB. In this figure, the x-axis represents the SHAP value, indicating the magnitude and direction (positive or negative) of each feature’s influence on the model’s prediction, while the y-axis lists features in descending order of importance. Each point reflects an individual respondent, with the color gradient from red to blue denoting the relative value of the feature (high to low, respectively). Higher values in family relationship satisfaction, health, goal clarity, financial safety, and personal safety (represented by red points on the right) were associated with higher predicted SWB. Loneliness showed a strong negative effect, with high values (red points) concentrated on the left. Return-home frequency and concern about public opinion had smaller but positive contributions.

**Fig. 4.**
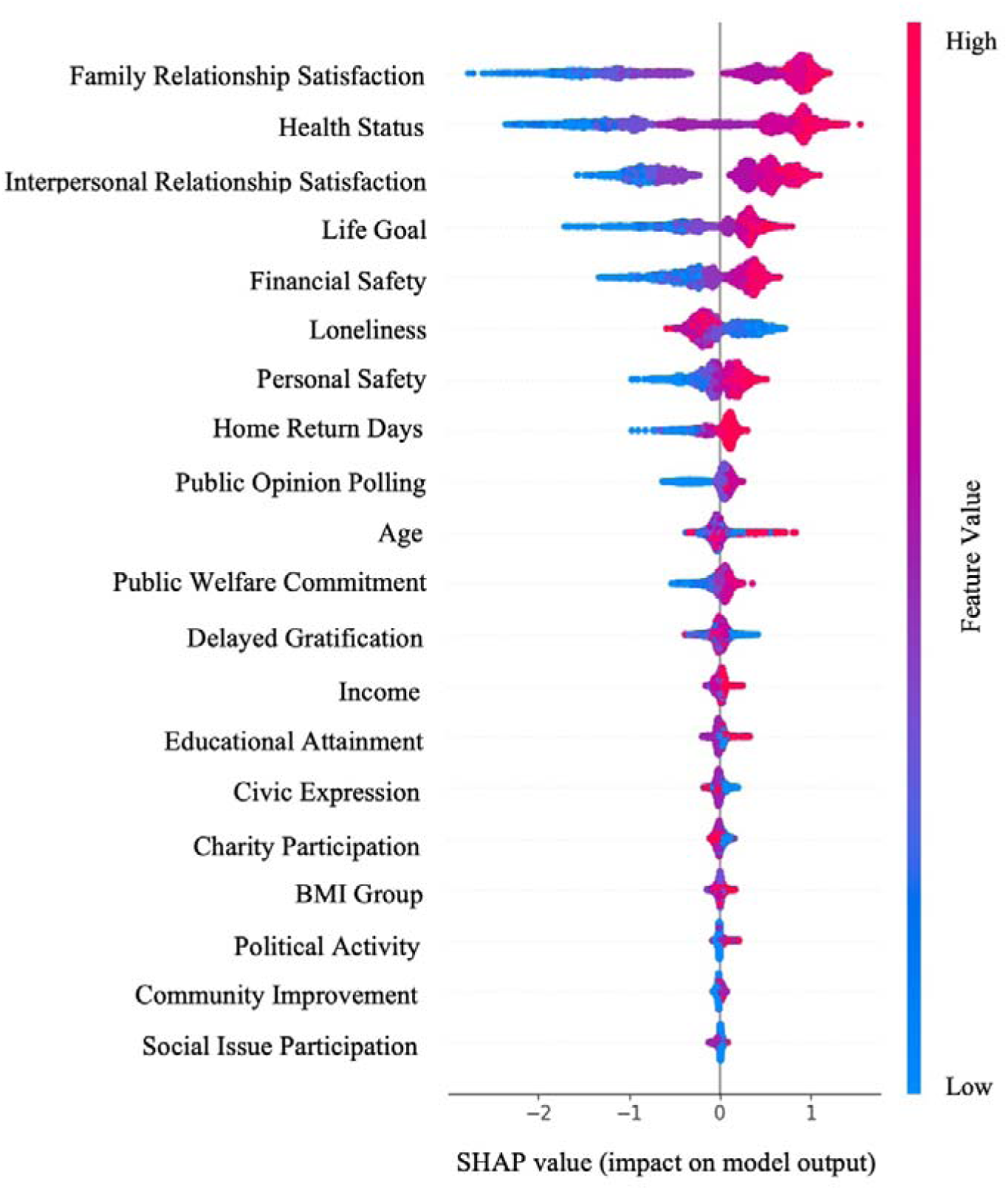
SHAP Summary Plot. The figure shows the distribution of SHAP values for the top features in the LightGBM model predicting subjective well-being. Each dot represents an individual prediction, with color indicating the feature value (red = high, blue = low). The direction of the SHAP value reflects the feature’s positive or negative contribution to well-being.

### Influence of Key Determinants and Threshold Changes on Subjective Well-Being

Dependence plots illustrate the presence of threshold effects and non-linear patterns *(see Fig. 5a–e).* This presentation approach allows us to explore each determinant’s threshold value, distinguish whether it functions as a protective or motivational factor, and examine the nonlinear relationship between determinants and SWB. As shown in *Fig. 5a–e*, family relationship satisfaction, interpersonal relationship satisfaction, and health status exhibit threshold effects, with limited influence on SWB at lower scores and a marked increase once mid-range values are exceeded. In contrast, life goal clarity and financial safety demonstrate a more linear and consistent association with SWB across the score range. Based on these patterns, the former set of variables can be considered protective factors, while the latter may be interpreted as motivational factors.

**Fig. 5a–e.**
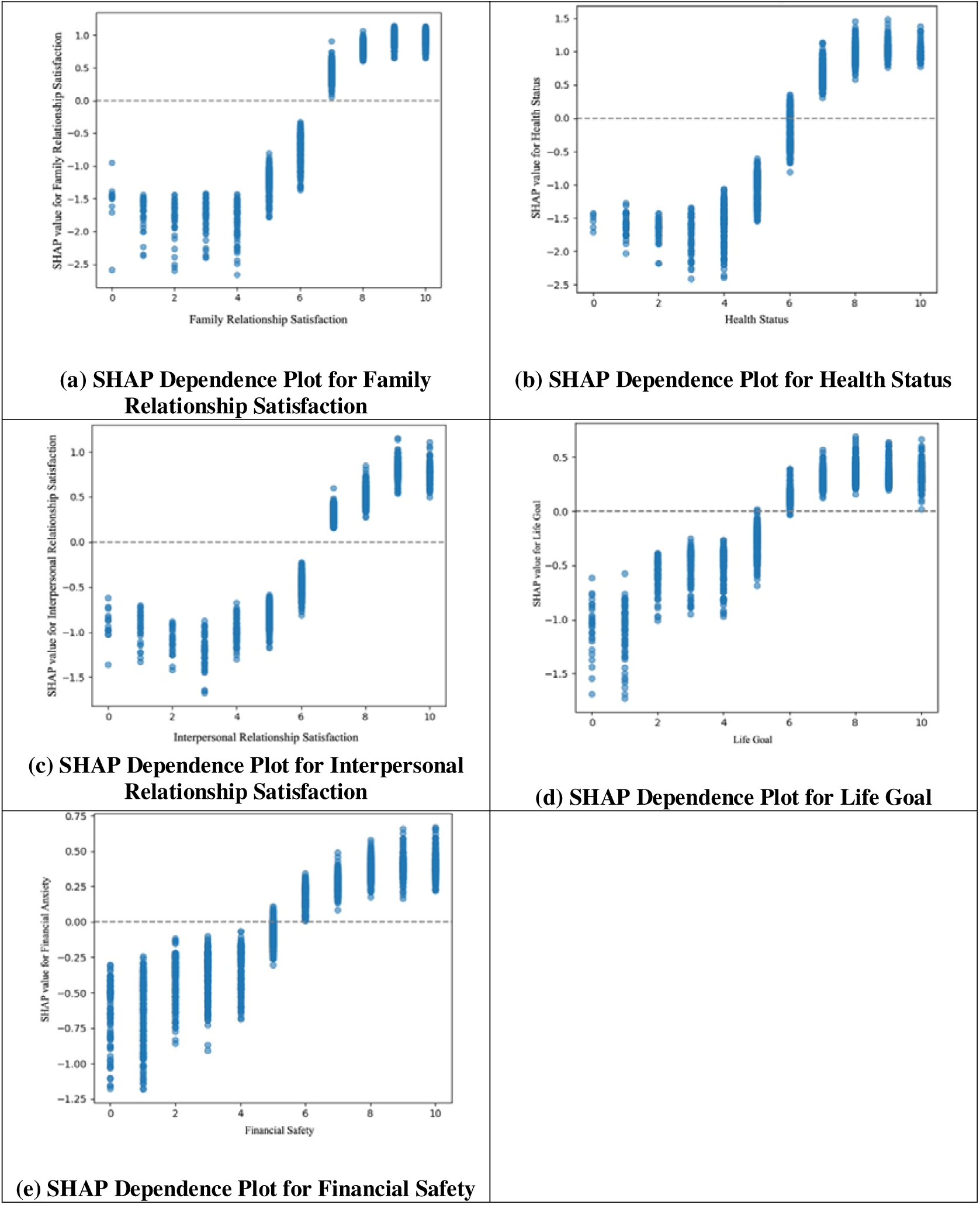
Dependence Plots for Top 5 Key Determinants (Original Scale) The figures display SHAP dependence plots for the five most influential features. Each plot shows the relationship between the original feature value (x-axis) and its SHAP value (y-axis), indicating the feature’s contribution to model predictions. Non-linear threshold effects are observed, with SHAP values rising notably only after scores exceed approximately 5 or 6 on the 0–10 scale.

### Sensitivity Analysis

To address potential bias in the dataset, we applied three different methods in the sensitivity analysis. The results from the sensitivity analyses were consistent with the main findings, suggesting that the results are robust *(see Appendices B, C, D)*.

## DISCUSSION

### Main Findings

This study offers a comprehensive and data-driven understanding of the key determinants of SWB in the Taiwanese adult population. Consistent with theoretical expectations, variables related to physical, psychological, and behavioral functioning collectively contributed nearly 70% of the model’s explanatory power. The five most influential predictors of SWB were, in order: family relationship satisfaction, interpersonal relationship satisfaction, self-rated health, clarity of life goal, and financial safety. The above factors appear to function through distinct pathways: financial safety and life goal clarity act as motivational drivers and exhibit linear associations with SWB, whereas family relationship satisfaction, interpersonal relationship satisfaction, and health status serve as protective buffers and display threshold effects, with SWB increasing markedly only after scores surpass a mid-range level (around 5 or 6 on a 10-point scale), indicating nonlinear relationships. Additionally, indicators such as loneliness, personal safety, and daily routine regularity (e.g., returning home before 10 p.m.) also emerged as meaningful contributors. In contrast to psychosocial and behavioral variables—which showed strong and often non-linear associations with SWB—commonly cited structural factors such as age, objective income, and educational attainment demonstrated limited predictive power. In terms of algorithmic performance, all evaluated models achieved acceptable predictive accuracy, with Gradient Boosting demonstrating the strongest and most consistent performance across accuracy and F1 score.

### Comparison with Existing Studies

In concrete terms, our study concludes five key SWB determinants—family relationship satisfaction, health status, interpersonal relationship satisfaction, life goals, and financial safety. These five factors align with the three main dimensions of Keyes’ model (Keyes, 2002), and our findings are also supported by local studies in Taiwan (Chang, 2009; Cheng et al., 2023; Lin et al., 2024). Our study also reveals that these key determinants can be categorized into protective and motivational factors, corresponding to different levels of Maslow’s hierarchy of needs. (Maslow, 1943). Specifically, stable family and interpersonal relationships offer emotional attachment and a sense of belonging, thereby strengthening social integration and highlighting the importance of social capital. Both international and local studies have pointed to the positive association between social capital and SWB (An et al., 2024; Chang, 2009; Wu, 2023). From a mental perspective, having a clear life goal not only provides individuals with direction and meaning, but also enhances their forward-looking mindset, which corresponds to the “Purpose and Meaning” dimension in VanderWeele’s theory of human flourishing (VanderWeele, 2017). Additionally, most of the key determinants exhibit a threshold effect. This means that once resource accumulation surpasses a certain threshold, SWB tends to grow rapidly rather than gradually. This phenomenon is aligned with the Theory of Fundamental Causes, which posits that structural social conditions influence well-being through the mechanisms of resource acquisition and accumulation (Link & Phelan, 1995). When critical resources reach a certain value, they may trigger a phase transition in well-being (Lynch et al., 2000; Ulm, 1991). This effect has been empirically validated by numerous international and local studies (Chung et al., 2021; Hsu et al., 2016; Lynch et al., 2000).

There remain several academic divergences regarding the conceptual framework of SWB, particularly concerning whether its dominant factors have universal applicability or if the model should be adjusted according to cultural characteristics and structural conditions. Our research partially supports the perspective of a cross-national model, since the local driving factors of SWB identified in our study correspond to the common determinants proposed by Diener et al (Diener et al., 2018). However, our research further reveals that the effects of these variables are mainly concentrated among well-being vulnerable groups. In these populations, the influence of these determinants appears stronger, suggesting that cultural context and structural position may play a moderating role. This finding aligns with the research of Oishi et al., which emphasizes that in the context of insufficient social or material support, individuals are more likely to rely on emotional support and safety-related interactive resources (Oishi et al., 1999). Under the Taiwan context, related research also indicates that relationship capital and the socioeconomic gap are the main factors driving well-being disparity, which is consistent with our findings (Chang, 2009; Yeh et al., 2015). Another notable academic argument concerns the relationship between income growth and SWB. The traditional view posits that economic advancement directly increases well-being, but the Easterlin Paradox challenges this assumption (Easterlin, 1974; Kahneman & Deaton, 2010). Our research reveals a result that provides a new explanation: although the connection between objective income and SWB is limited, perceived financial safety shows a clear linear contribution to well-being. When individuals feel higher financial safety, their well-being increases steadily, without evidence of diminishing marginal returns. Hence, in public policy design, rather than focusing narrowly on improving objective income levels, priority should be given to addressing relative deprivation and financial unsafety. This finding is consistent with the systematic review by Ngamaba et al. and also echoes some local studies (Kuo et al., 2024; Ngamaba et al., 2017), which suggest that subjective personal perceptions and equity expectations should be central considerations.

### Policy implication

The study revealed actionable policy implications grounded in both global frameworks and Taiwan’s specific context. The World Health Organization (WHO) and Organisation for Economic Co-operation and Development (OECD) have consistently emphasized that without social connection, individual health status, financial safety, and life goal clarity, people may become part of the SWB vulnerable group, which can lead to poor physical and mental health (Mahoney et al., 2024; OECD, 2024a, 2024b; World Health Organization, 2025). OECD further emphasizes that improvements in SWB measurement must meet a certain threshold to generate meaningful effects (OECD, 2024b). In line with these priorities, WHO, OECD, and the UN recommend the following policies: First, governments should implement community development programs to enhance social cohesion, such as creating multifunctional social spaces and organizing activities like family interaction days (OECD, 2013, 2020). Second, governments should develop action plans to foster health behavior, evaluate policy feedback, and organize health promotion activities (World Health Organization, 2021). Third, governments should strengthen the social security network, improve policies related to social welfare coverage, and increase residents’ financial literacy through training programs to reduce economic stress (OECD, 2024a; Osberg, 2021). In education and employment-focused empowerment programs, schools should provide career counseling to cultivate intrinsic motivation and support mid- and late-life career restructuring (Abdallah, 2024). Furthermore, OECD highlights that setting a well-being baseline allows for resource targeting toward low-SWB groups and enables stratified policy interventions through public space engagement, helping people cross the critical threshold to improved well-being (OECD, 2013). After implementation, a cost-effectiveness analysis should be conducted, integrating the results of threshold effects. Our findings support international priorities by demonstrating that the factors identified above positively contribute to SWB. Evidence indicates that optimizing these determinants can enhance individual well-being and generate secondary benefits, including improved health and strengthened social bonds. Therefore, when governments formulate public policy, priority should be given to strengthening social networks, promoting individual health, clarifying life direction, and preventing poverty risks.

Regarding efforts in Taiwan, the Ministry of Education has implemented the Third Phase of the Mid-term Family Education Promotion Program to enhance family interaction through family growth centers and counseling services (Ministry of Education, 2025a). In alignment with WHO’s emphasis on the health benefits of social integration, the government has concurrently promoted the Community Empowerment Project, organizing local events and encouraging resident participation to foster mutual support (Tsai, 2014) . Moreover, our study suggests that governments should design region-specific interventions for resource-disadvantaged areas to improve SWB, for instance, using volunteer systems to reinforce social structure density and individual well-being (Chang, 2009). In the health promotion domain, long-term policy efforts have led to the establishment of Health Promoting Institutions across most healthcare facilities, supporting organizational restructuring, implementing early cancer detection protocols, forming health advocacy teams, and reinforcing the service network through regular audits and certification processes (Ministry of Health and Welfare, 2015). To support life goal development, the government also promotes the Mid-term Plan for Life Education Program, actively encouraging schools to implement life education activities that guide students in building identity awareness (Ministry of Education, 2022). For middle-aged and older populations, the government has launched the Pilot Project for the Third Life University Program, which expands learning opportunities and supports reeducation to help individuals pursue unfinished goals and strengthen their sense of life purpose (Ministry of Education, 2025b). In the future, governments may expand free course resources for vulnerable middle-aged and older adults to help them reframe their life goals. Regionally, the Happy Station Project, developed by local authorities in partnership with convenience store operators, serves as a creative model. It establishes a community-based real-time care system that offers basic emergency support to children and adolescents, facilitates referrals to support institutions, and builds a resilient regional safety net (New Taipei City Government, 2025). Beyond traditional income support or workplace counseling to raise earnings, and in response to the threshold effect of SWB, our research suggests that the government should consider launching an "investment-oriented well-being promotion pilot program." This program would integrate Pay-for-Performance mechanisms from the health insurance system with person-centered investment interventions (Gonzalez-Arcos et al., 2023). The program could target low-income individuals whose SWB falls below a specific threshold, using community-based applications. After review, the government would provide small incentive subsidies. If participants successfully cross the well-being threshold, additional performance-based rewards could be issued. This approach would promote grassroots involvement in well-being enhancement (Gonzalez-Arcos et al., 2023). The program reflects the non-homogeneous nature of well-being improvement and offers a potential public return on investment with low cost and high impact.

### Research Limitations

There are several caveats to this study. First, during the preprocessing stage, the treatment of outliers and the deletion of high-dominant features may have led to the loss of core information, potentially affecting the comprehensiveness and explanatory power of the research findings. Second, when applying class balancing techniques, the use of synthetic samples generated by SMOTE may have introduced distributional shifts, which could impact the generalizability of the results. Third, although machine learning models such as XGBoost and LightGBM effectively enhance predictive performance, their complex model structures make it difficult to clearly explain the relationships between features and outcomes. This complexity may limit the operational feasibility and visibility of the findings for policy design and practical application. Finally, the dataset did not include options beyond the traditional binary gender categories. Specifically, sex was categorized as male or female based on self-reported responses. The questionnaire did not explicitly specify whether the reported sex referred to biological sex or gender identity., which may limit the generalizability of the findings to gender-diverse populations.

## CONCLUSION

This study identifies several actionable levers for enhancing SWB, including strengthening social relationships, improving health status, clarifying life goals, and ensuring financial safety. Importantly, our findings emphasize the presence of threshold effects, suggesting that the benefits of intervention are not uniformly distributed but are particularly pronounced among individuals starting from lower baseline conditions—often those most vulnerable. These insights align closely with multiple SDGs, particularly SDG 3 (Good Health and Well-being), as well as goals related to reducing inequality, promoting social resilience, supporting educational and economic empowerment, and preventing poverty. The identification of motivational versus protective factors also provides a nuanced framework for prioritizing intervention strategies across different population segments.

## Data Availability

The dataset used in this study (year 2024) is not currently available for public access, as it is undergoing curation and approval processes. Earlier waves of the dataset are publicly available through Academia Sinica at https://srda.sinica.edu.tw/plan/?idx=SRDA.AS039. The 2024 wave will be released for download in the future once data curation and approval are completed.

## DECLARATIONS

### Author Contributions

Yeh, Kuo, Hsieh, Lu, and Lee contributed to the conceptualization and methodology of the study. Formal analysis was conducted by Yeh. Lee provided supervision. The original draft was prepared by Yeh, and all authors (Yeh, Kuo, Hsieh, Lu, and Lee) contributed to the review and editing of the manuscript.

### Ethical Statement

This study was approved by the Institutional Review Board for Humanities & Social Science Research at Academia Sinica (AS-IRB-HS 02-19025(R15)) and the Research Ethics Committee of National Taiwan University (No. 202411HS043). The survey data were retrospectively de-identified to ensure compliance with ethical research guidelines. The data analysis was conducted after de-identification, in order to safeguard the privacy of the respondents and uphold data confidentiality principles.

### Consent

For the questionnaire data used in this study, all respondents signed the informed consent form prior to participation.

### Code Availability

Statistical analysis code in Python (version 3.12.2) and Stata (version 18) is available for download at: https://github.com/johntayuleeHEPI/well-being_machine-learning/tree/main.

## Appendices A. Detailed Results Supporting Main Text

**Table A1.**
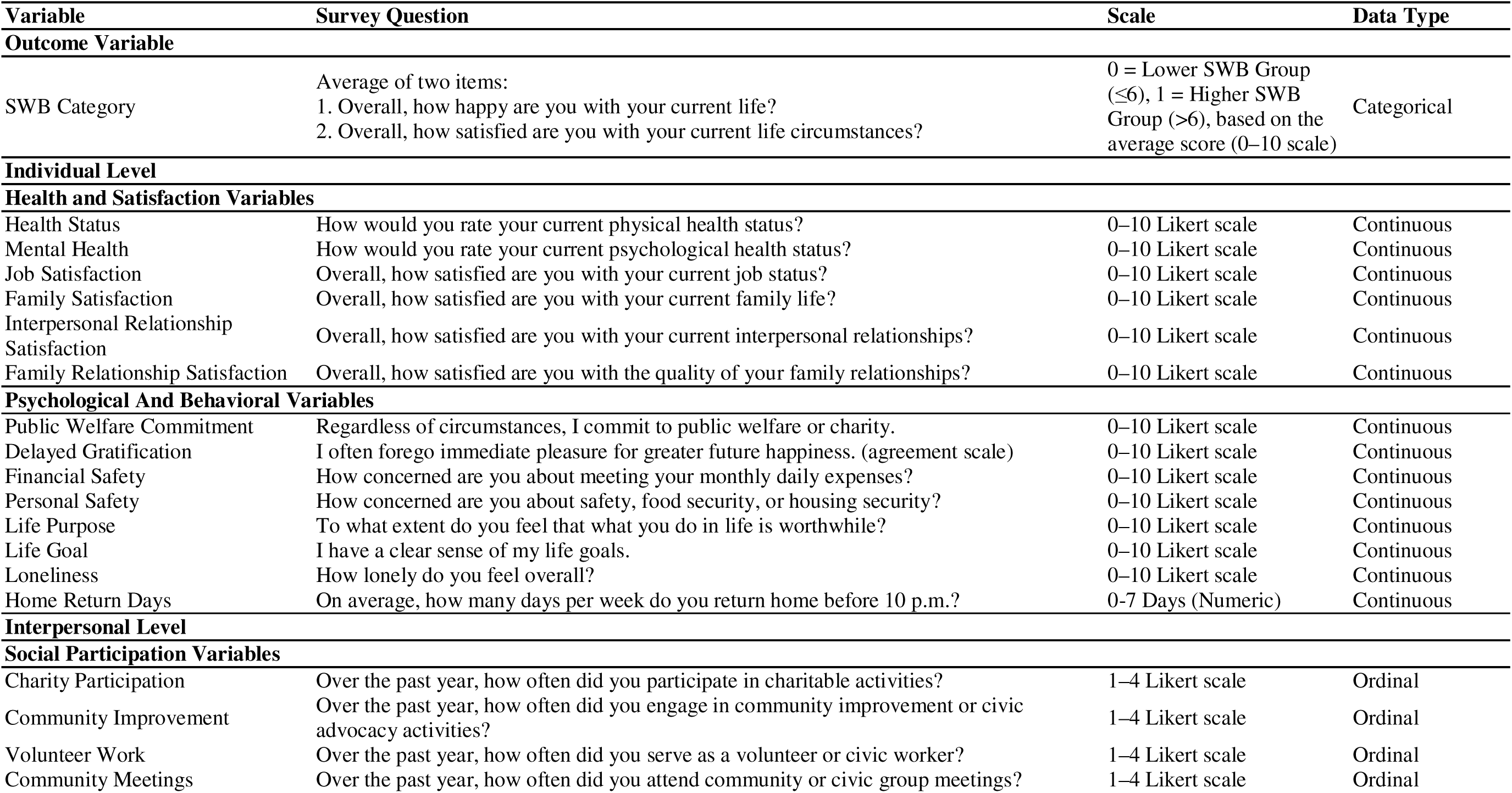
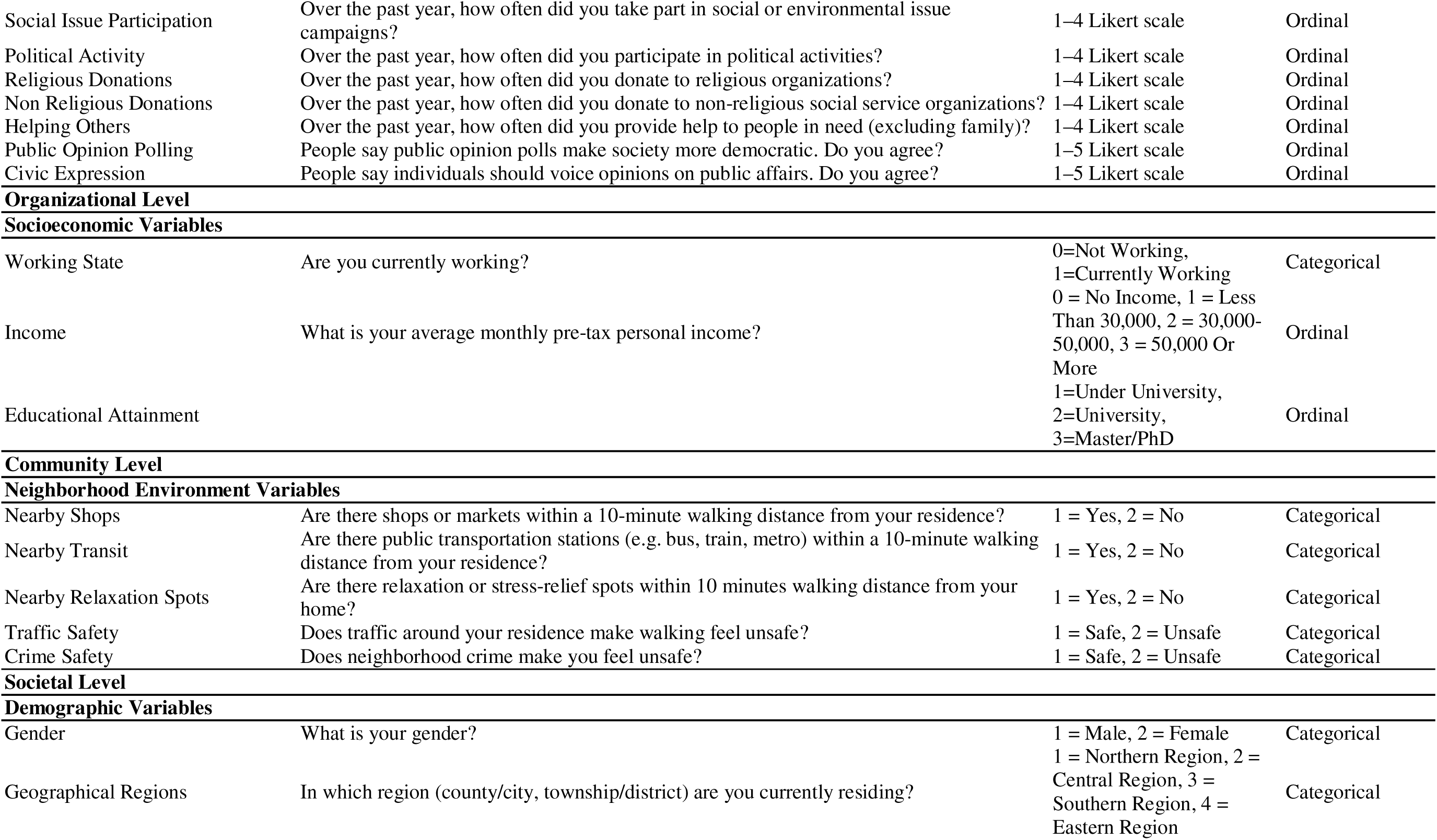
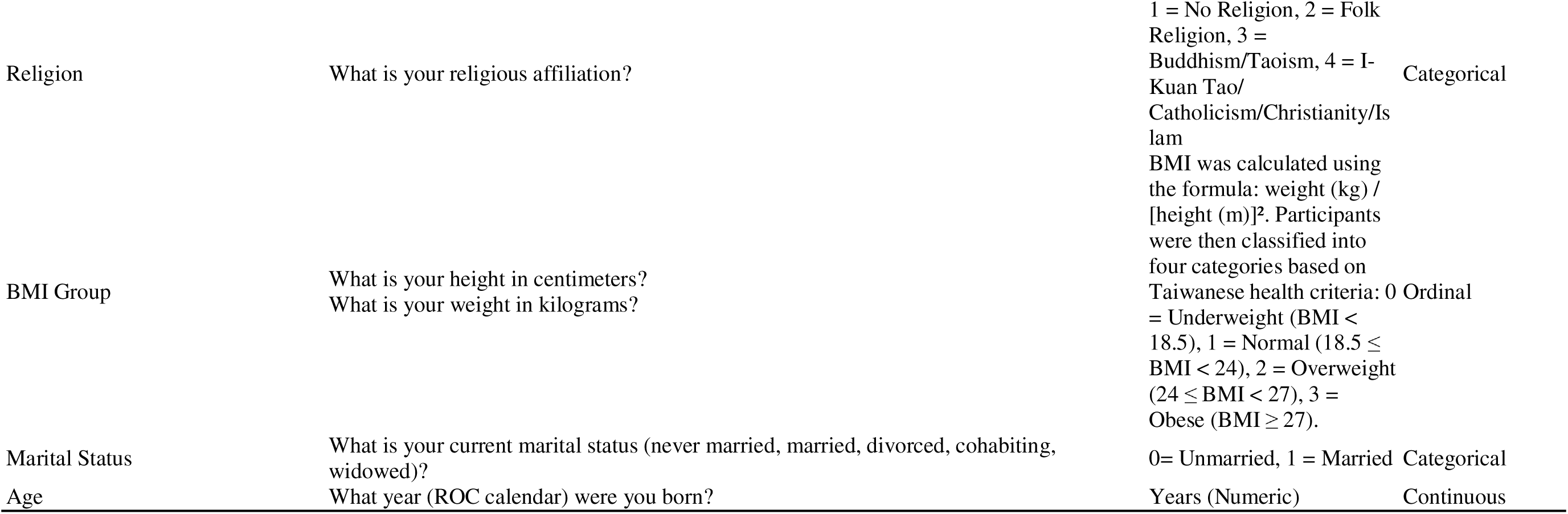
Definitions and Descriptions of Variables Classified According to the Social Ecological Model.

**Fig. A1.**
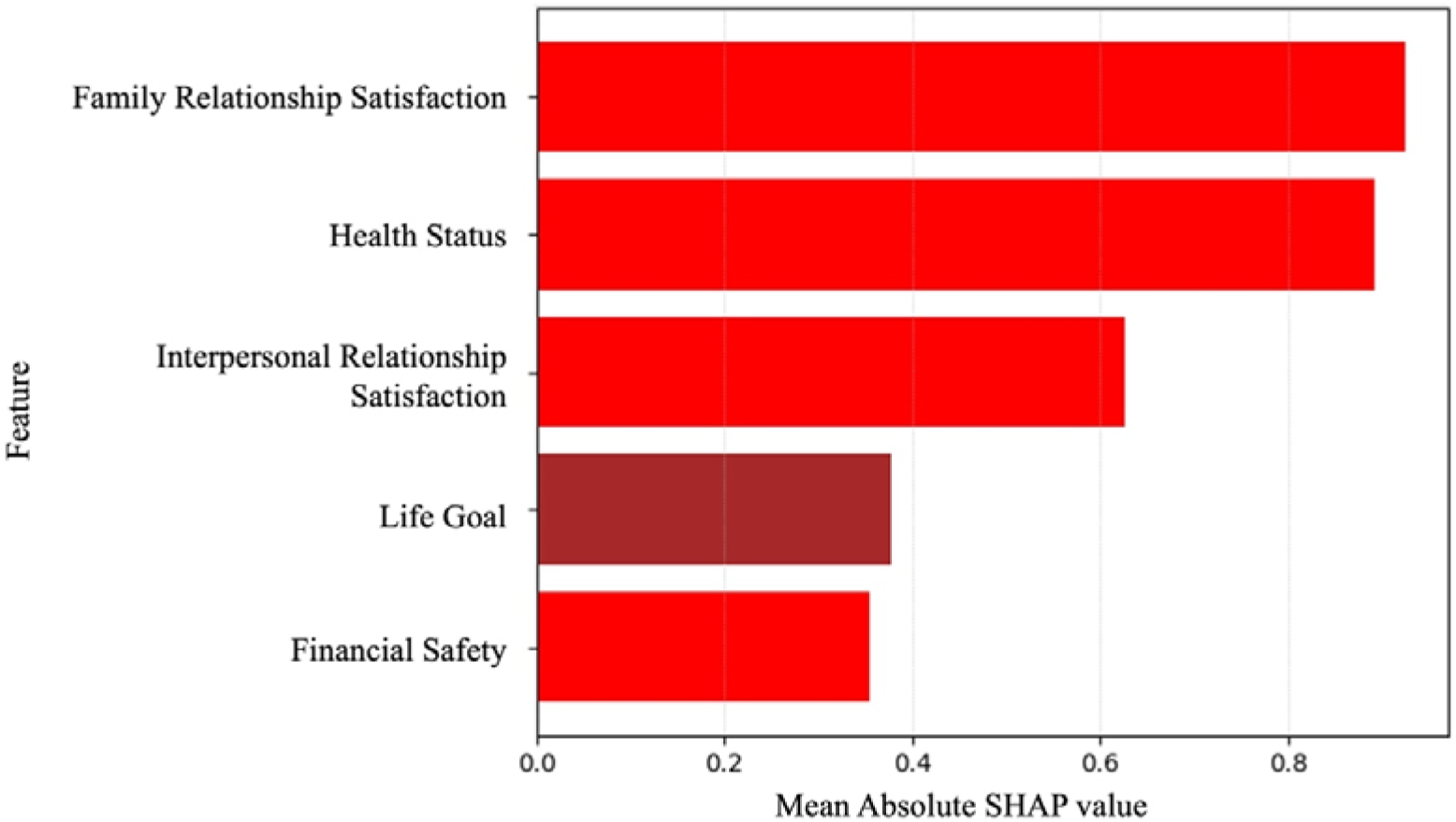
Top 5 Features by Aggregated SHAP Value. The figure shows the five most influential predictors of subjective well-being based on mean absolute SHAP values from the LightGBM model. Family relationship satisfaction and health status exhibit the strongest contributions, followed by interpersonal relationship satisfaction, life goal clarity, and financial anxiety.

**Fig. A2a–e.**
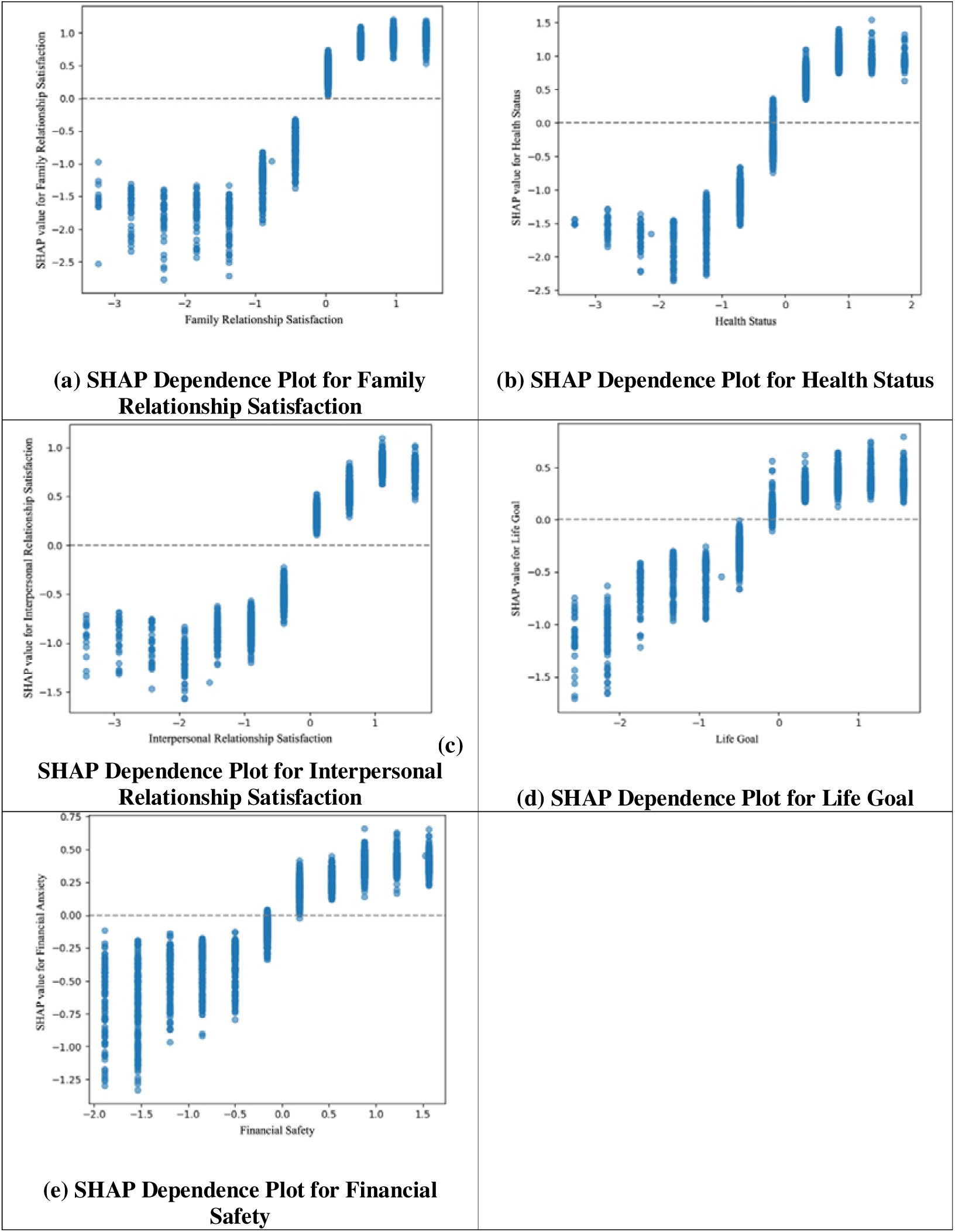
Dependence Plots for Top 5 Key Determinants (Standardized Scale) The figures present SHAP dependence plots for the five most influential features of subjective well-being, using standardized scores. Each plot illustrates the relationship between the standardized feature value and its SHAP value, reflecting the feature’s contribution to model predictions.

## Appendices B. Sensitivity Analyses: SWB as a Continuous Variable

**Table B1.** Model Performance Metrics.

| Model | R Square | MAE | RMSE |
| --- | --- | --- | --- |
| LinearRegression | 0.659 | 0.866 | 1.215 |
| Lasso | 0.648 | 0.887 | 1.234 |
| Ridge | 0.659 | 0.866 | 1.215 |
| LightGBM | 0.666 | 0.856 | 1.201 |

**Fig. B1.**
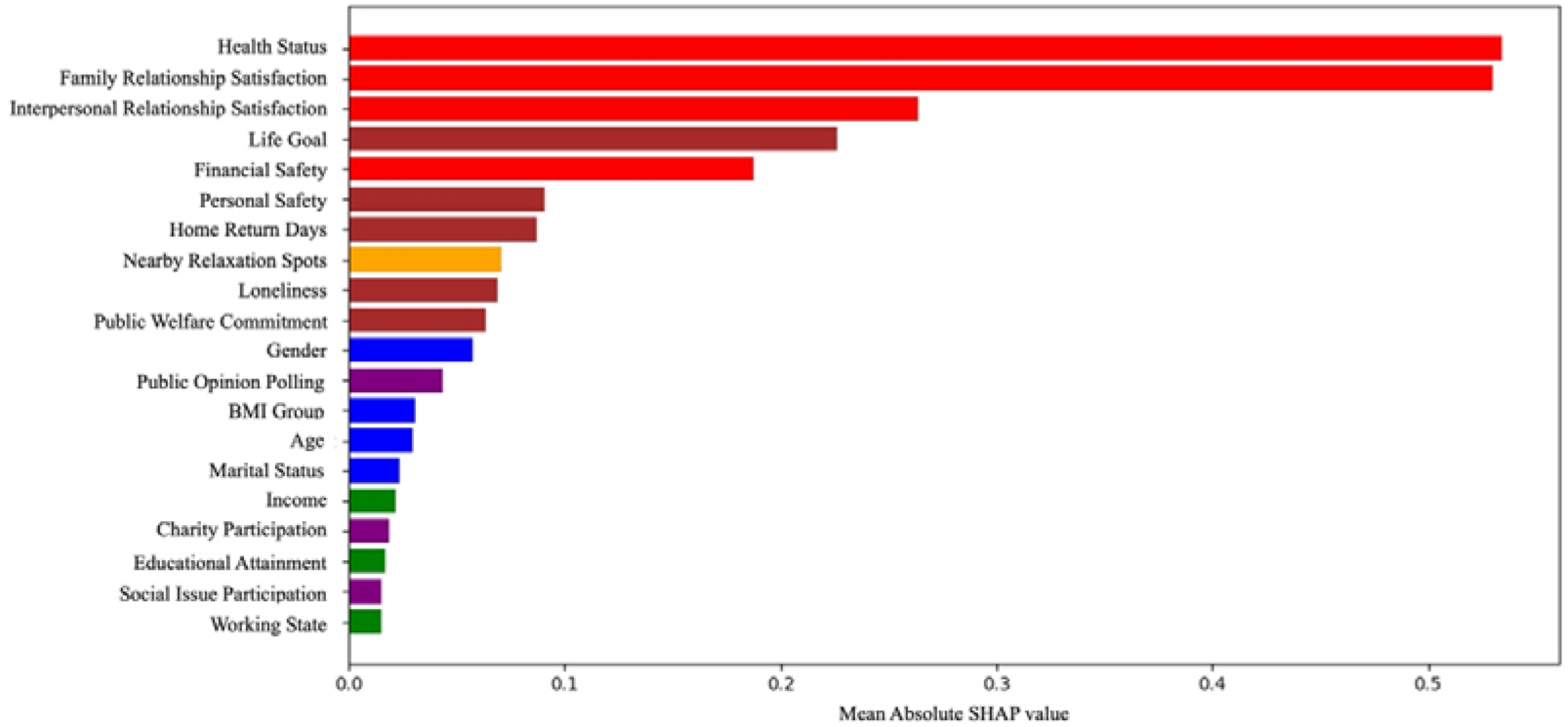
Top Features by Aggregated SHAP Value. The figure shows the top predictors of subjective well-being based on SHAP values from the LightGBM model. Higher SHAP values reflect greater model contribution. Variables are color-coded by domain: health/satisfaction (red), psychological/behavioral (brown), socioeconomic (green), demographic (blue), neighborhood (orange), and social participation (purple).

**Fig. B2.**
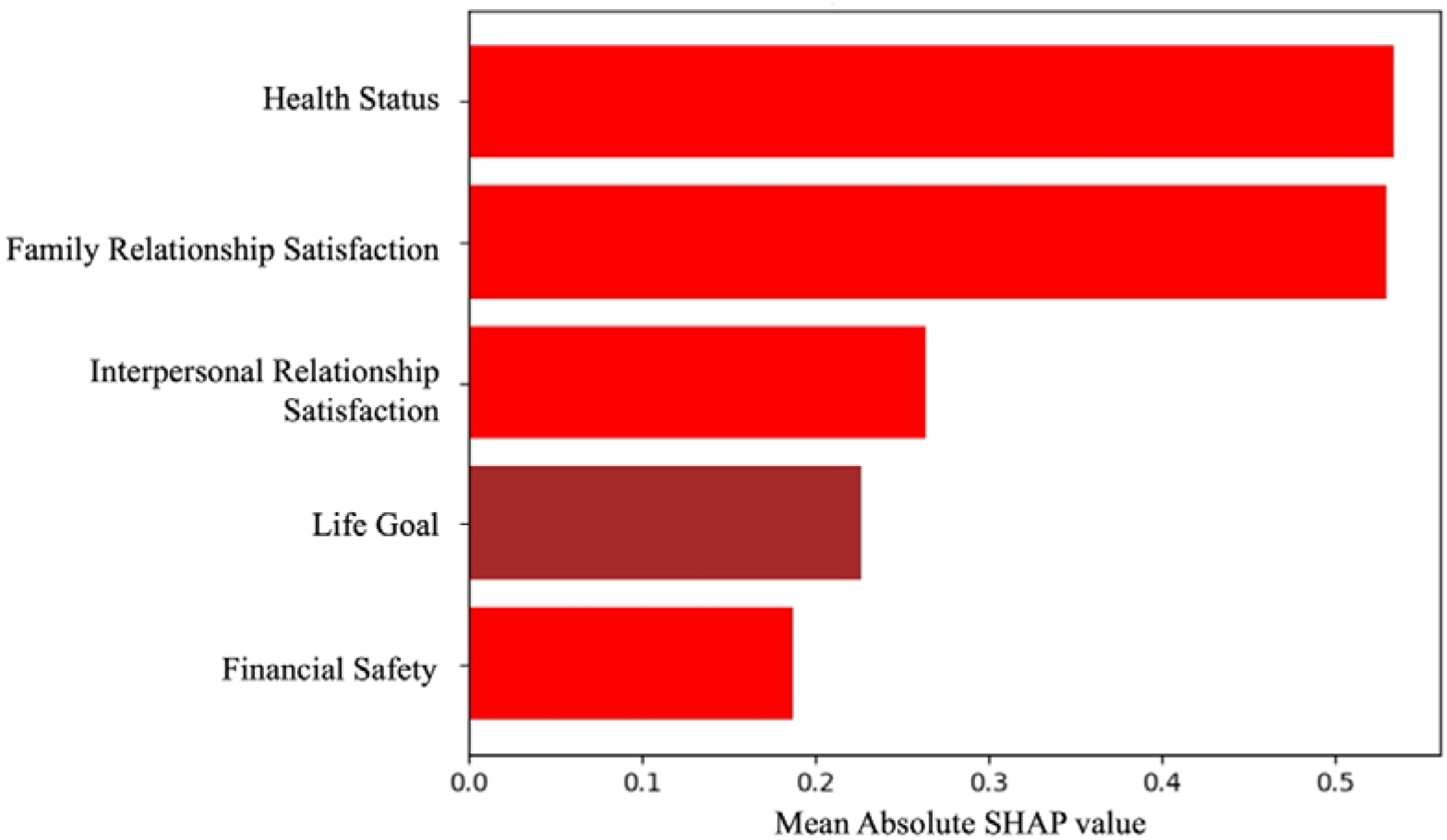
Top 5 Features by Aggregated SHAP Value. The figure shows the five most influential predictors of subjective well-being based on mean absolute SHAP values from the LightGBM model. Family relationship satisfaction and health status exhibit the strongest contributions, followed by interpersonal relationship satisfaction, life goal clarity, and financial anxiety.

**Fig. B3.**
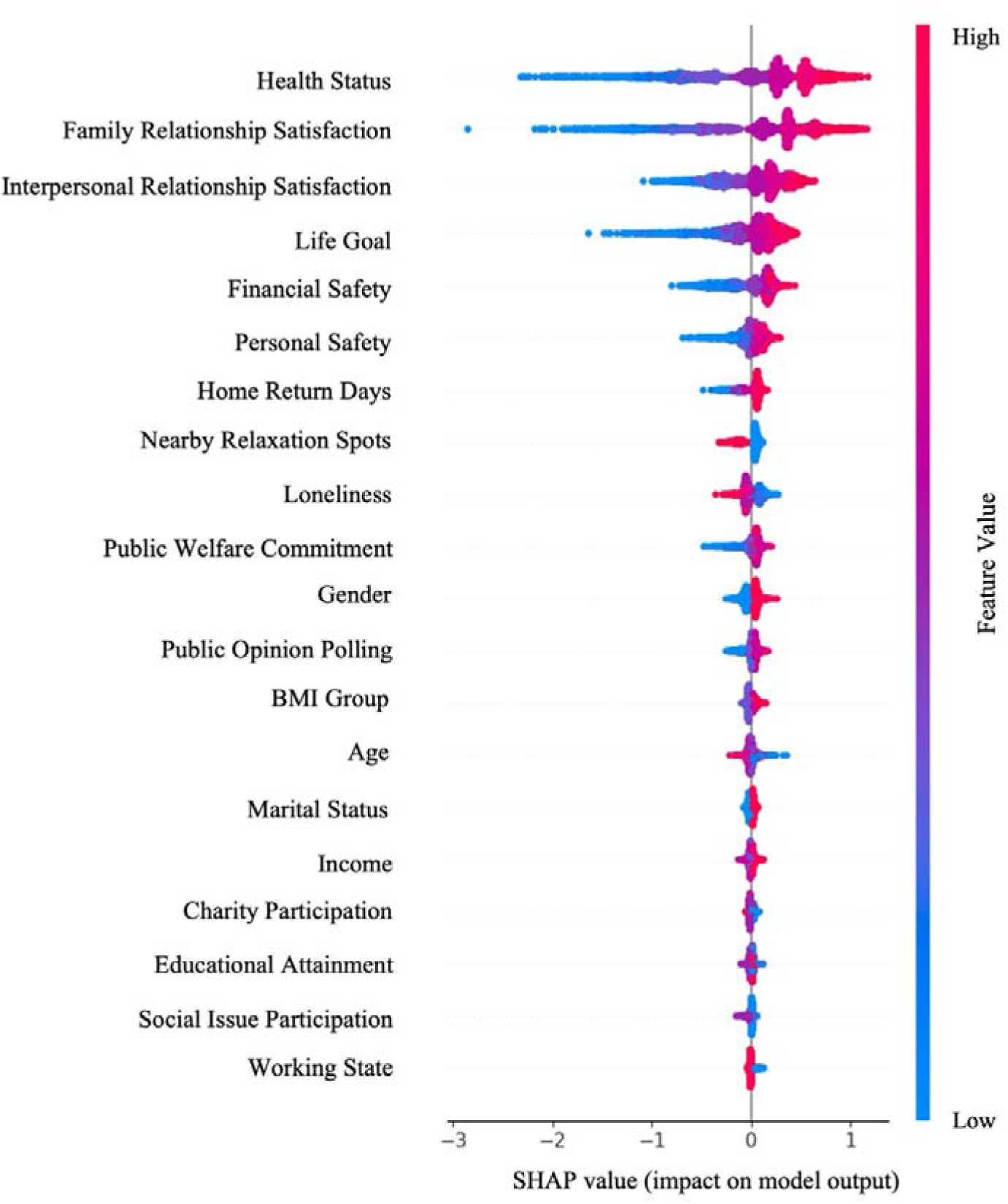
SHAP Summary Plot. The figure shows the distribution of SHAP values for the top features in the LightGBM model predicting subjective well-being. Each dot represents an individual prediction, with color indicating the feature value (red = high, blue = low). The direction of the SHAP value reflects the feature’s positive or negative contribution to well-being.

**Fig. B4a–e.**
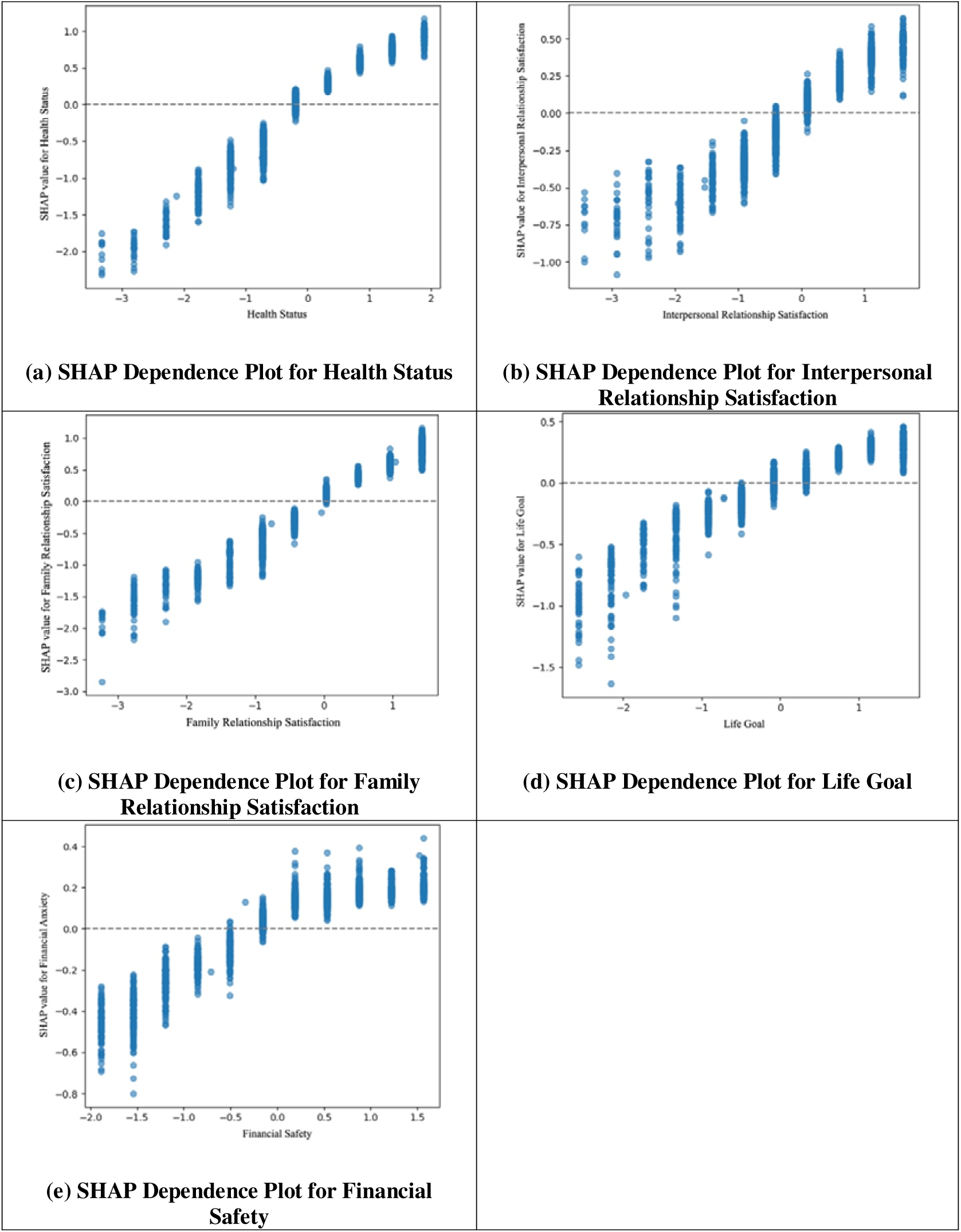
Dependence Plots for Top 5 Determinants. The figures present SHAP dependence plots for the five most influential features of subjective well-being, using standardized scores. Each plot illustrates the relationship between the standardized feature value and its SHAP value, reflecting the feature’s contribution to model predictions.

## Appendices C. Sensitivity Analyses: SWB Dichotomized at 7-Point Cutoff

**Table C1.** Model Evaluation and Predictive Performance of Machine Learning Models.

| Model | Accuracy | Sensitivity | Specificity | Precision | F1 Score | AUROC |
| --- | --- | --- | --- | --- | --- | --- |
| Logistic | 0.830 | 0.799 | 0.849 | 0.757 | 0.778 | 0.908 |
| Random Forest | 0.826 | 0.770 | 0.860 | 0.764 | 0.767 | 0.909 |
| Gradient Boosting | 0.827 | 0.778 | 0.857 | 0.762 | 0.770 | 0.911 |
| XGBoost | 0.829 | 0.774 | 0.862 | 0.768 | 0.771 | 0.911 |
| LightGBM | 0.823 | 0.764 | 0.857 | 0.760 | 0.762 | 0.910 |

**Fig. C1.**
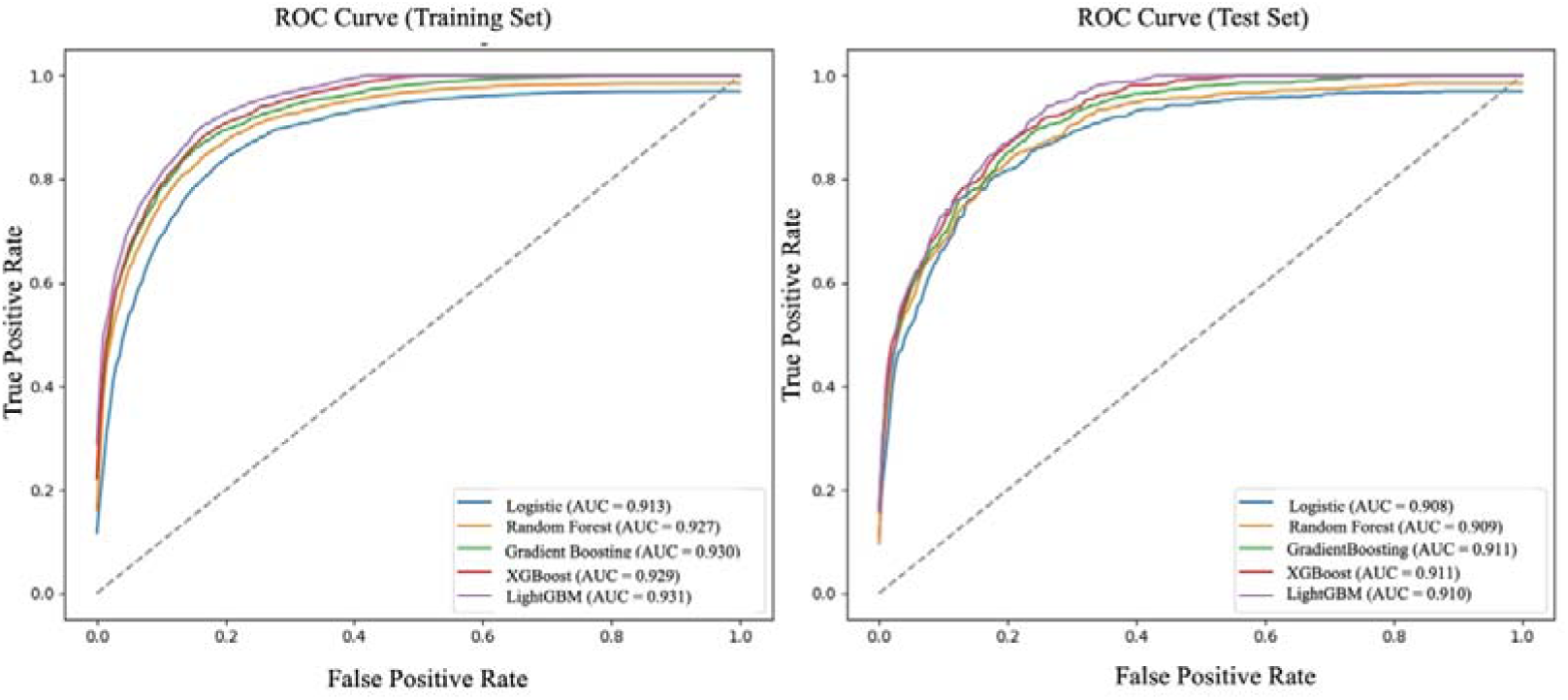
ROC Curves for Training and Testing Sets. The figure presents Receiver Operating Characteristic curves for five machine learning models. The left panel shows ROC curves for the training set, and the right panel for the test set. Gradient Boosting and XGBoost achieved the highest Area Under the Curve values.

**Fig. C2.**
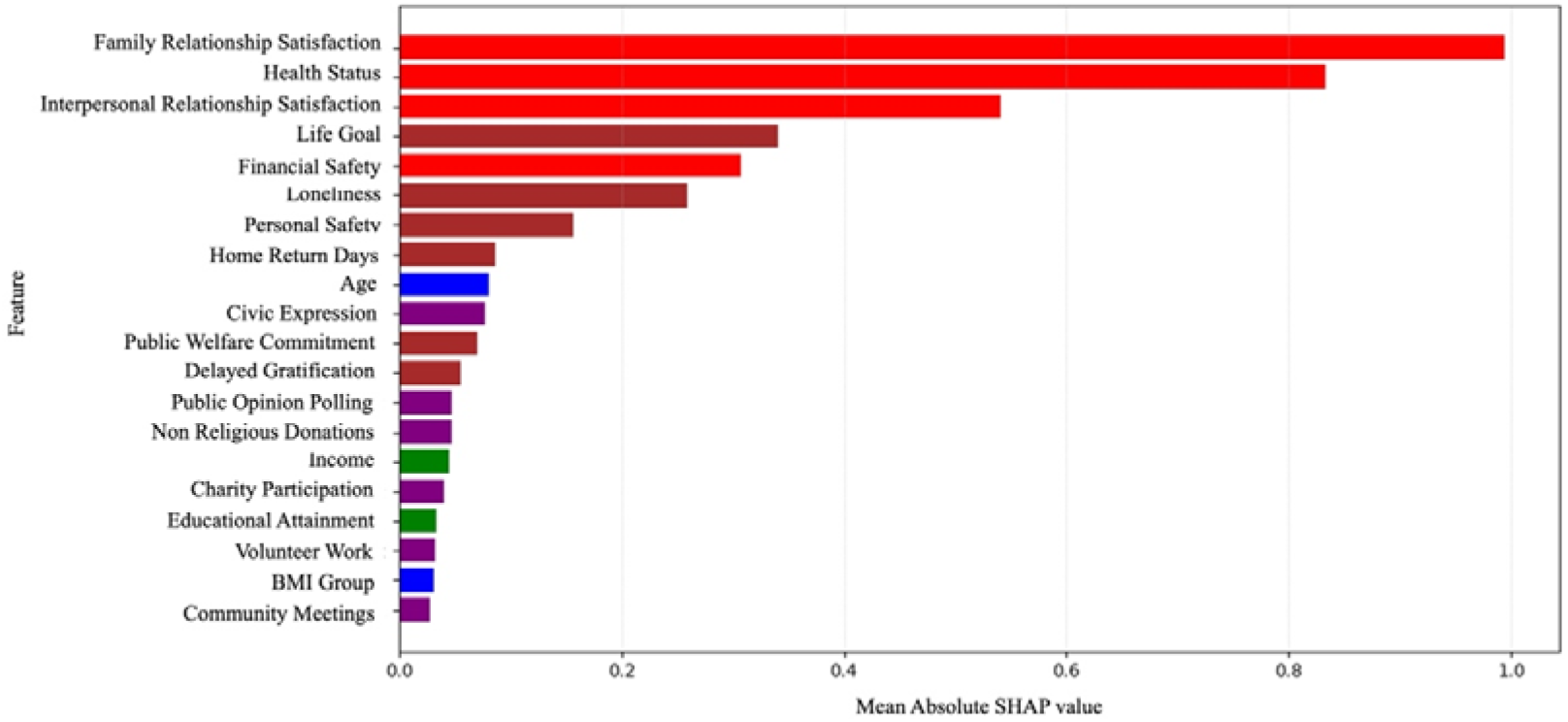
Top Features by Aggregated SHAP Value. The figure shows the top predictors of subjective well-being based on SHAP values from the LightGBM model. Higher SHAP values reflect greater model contribution. Variables are color-coded by domain: health/satisfaction (red), psychological/behavioral (brown), socioeconomic (green), demographic

**Fig. C3.**
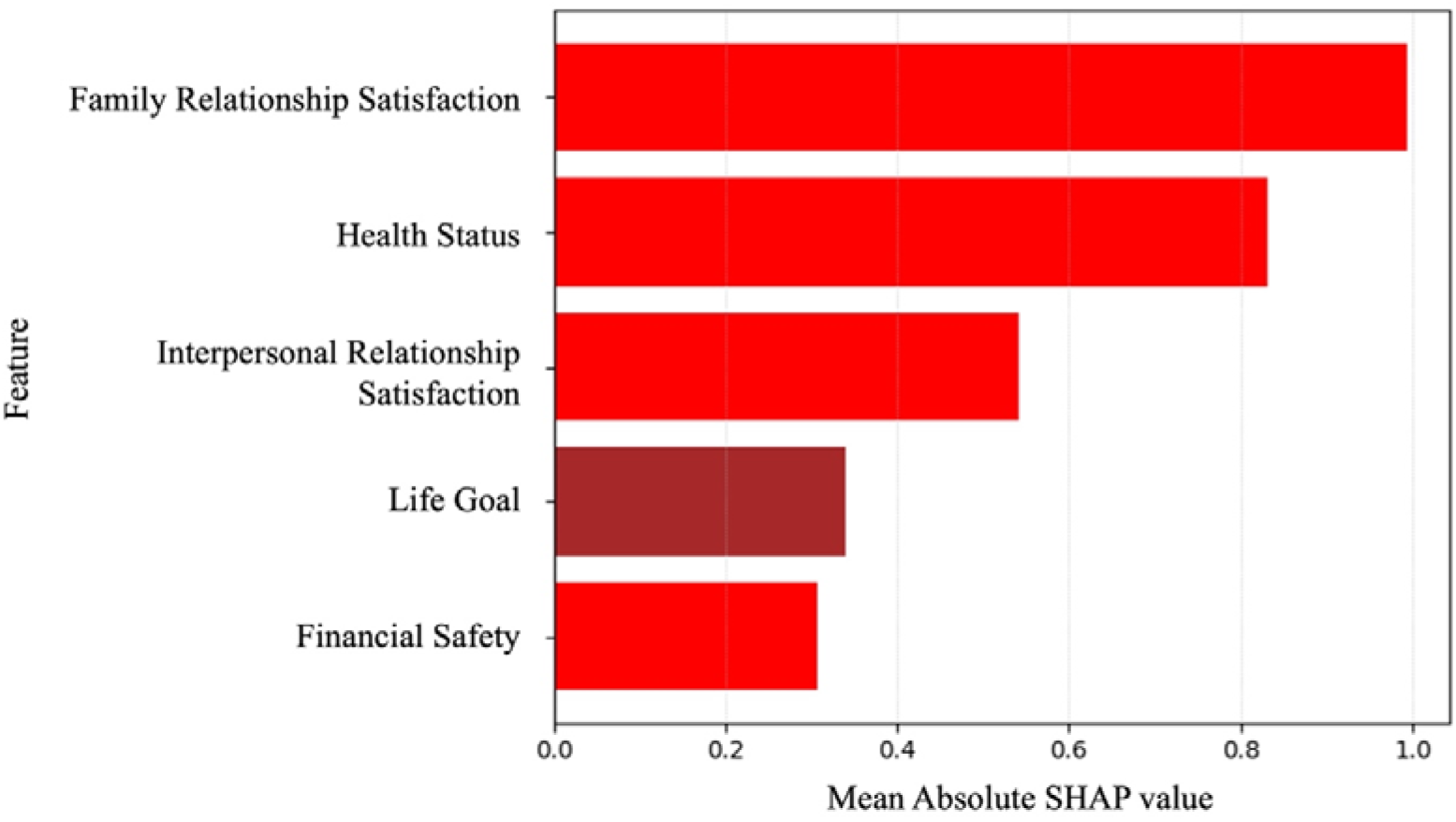
Top 5 Features by Aggregated SHAP Value. The figure shows the five most influential predictors of subjective well-being based on mean absolute SHAP values from the LightGBM model. Family relationship satisfaction and health status exhibit the strongest contributions, followed by interpersonal relationship satisfaction, life goal clarity, and financial anxiety.

**Fig. C4.**
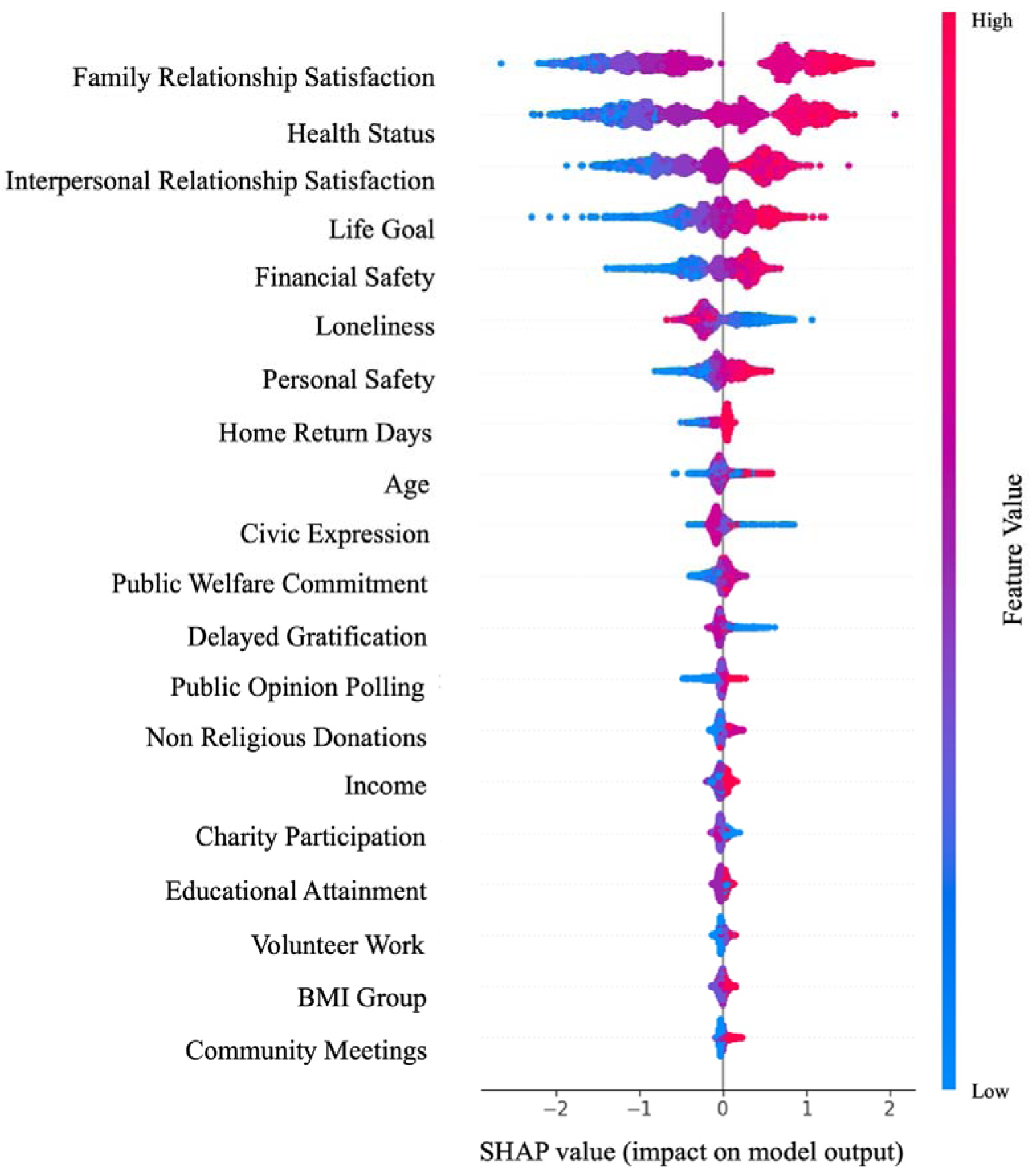
SHAP Summary Plot. The figure shows the distribution of SHAP values for the top features in the LightGBM model predicting subjective well-being. Each dot represents an individual prediction, with color indicating the feature value (red = high, blue = low). The direction of the SHAP value reflects the feature’s positive or negative contribution to well-being.

**Fig. C5a–e.**
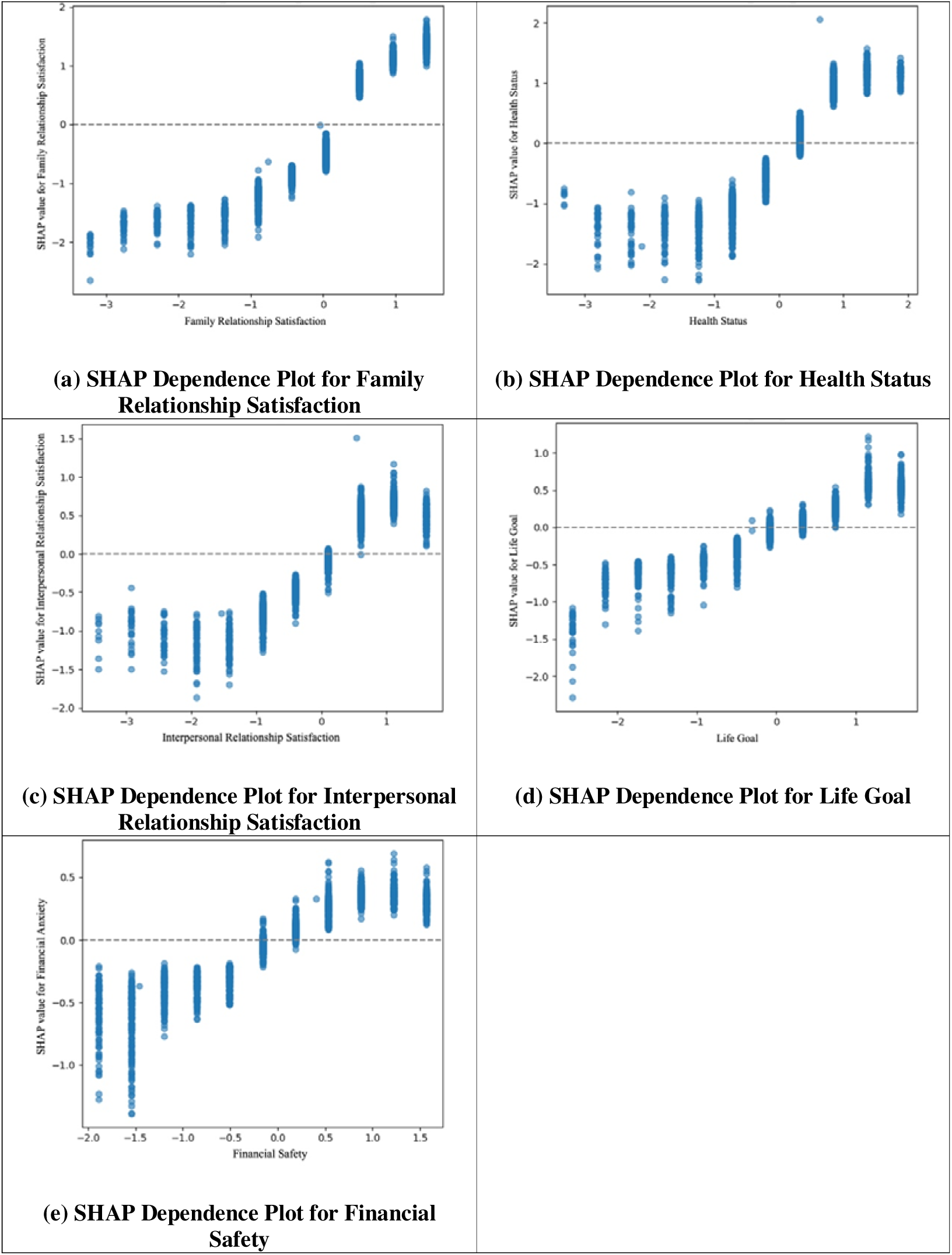
Dependence Plots for Top 5 Determinants. The figures present SHAP dependence plots for the five most influential features of subjective well-being, using standardized scores. Each plot illustrates the relationship between the standardized feature value and its SHAP value, reflecting the feature’s contribution to model predictions.

## Appendices D. Sensitivity Analyses: Subgroup Fairness Analyses

**Table D1.** Model Evaluation and Predictive Performance of Machine Learning Models.

| Model | Sensitive Attributes | Accuracy | AUROC | F1 Score | Sensitivity | Specificity |
| --- | --- | --- | --- | --- | --- | --- |
| Logistic | Gender | 0.835 | 0.908 | 0.784 | 0.808 | 0.851 |
| Logistic | Income | 0.829 | 0.907 | 0.778 | 0.809 | 0.840 |
| Random Forest | Gender | 0.830 | 0.907 | 0.768 | 0.758 | 0.872 |
| Random Forest | Income | 0.823 | 0.908 | 0.756 | 0.741 | 0.871 |
| Gradient Boosting | Gender | 0.826 | 0.911 | 0.767 | 0.771 | 0.858 |
| Gradient Boosting | Income | 0.826 | 0.910 | 0.769 | 0.779 | 0.854 |
| XGBoost | Gender | 0.820 | 0.895 | 0.757 | 0.756 | 0.857 |
| XGBoost | Income | 0.813 | 0.901 | 0.748 | 0.747 | 0.852 |
| LightGBM | Gender | 0.825 | 0.911 | 0.762 | 0.755 | 0.866 |
| LightGBM | Income | 0.823 | 0.909 | 0.765 | 0.778 | 0.849 |

**Table D2.**
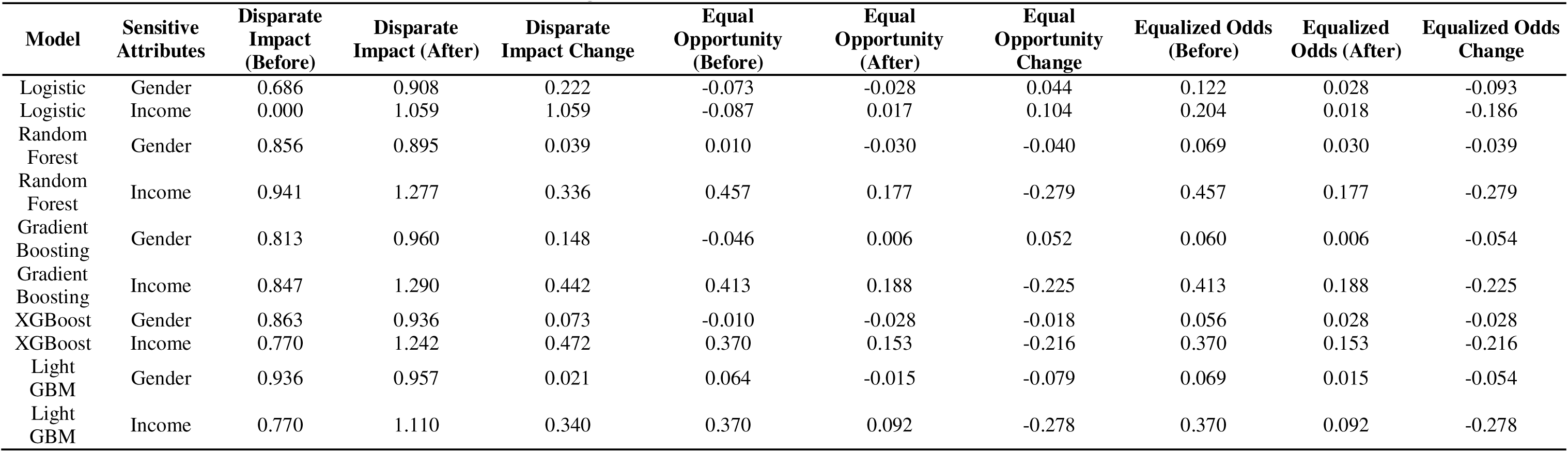
Model Evaluation and Predictive Performance of Machine Learning Models.

**Fig. D1.**
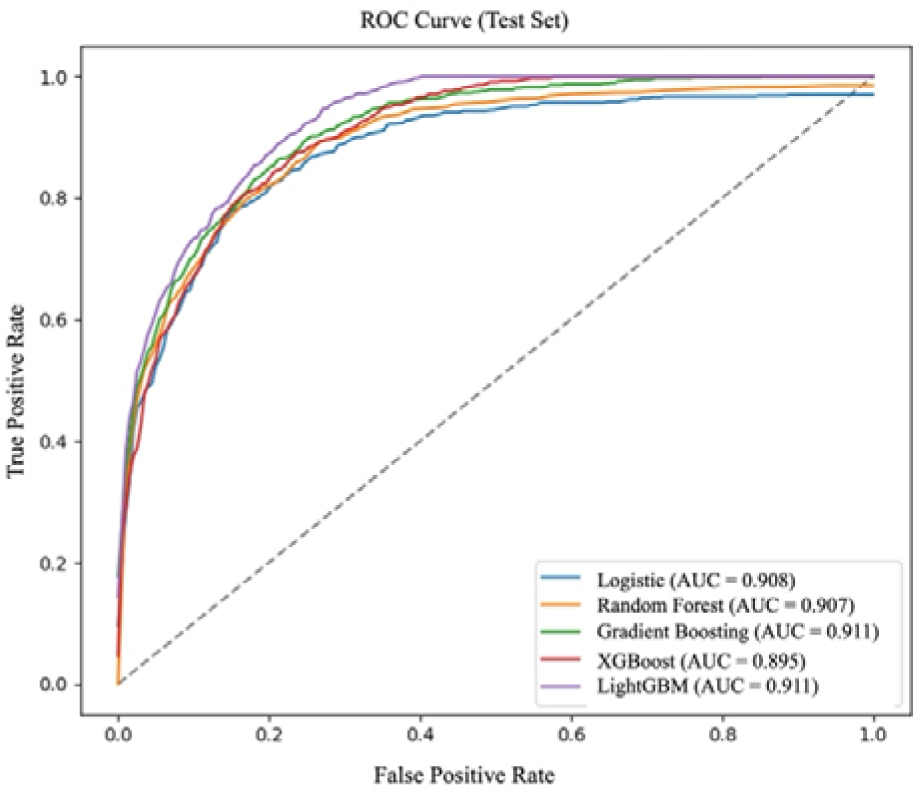
ROC Curves for Subgroup Fairness Models (Gender as Sensitive Attribute) The figure presents ROC curves for models incorporating subgroup fairness analysis using gender as the sensitive attribute. All models achieved high AUC values (ranging from 0.895 to 0.911), indicating strong classification performance across gender subgroups. This suggests that predictive accuracy remained consistent while accounting for potential gender-based disparities.

**Fig. D2.**
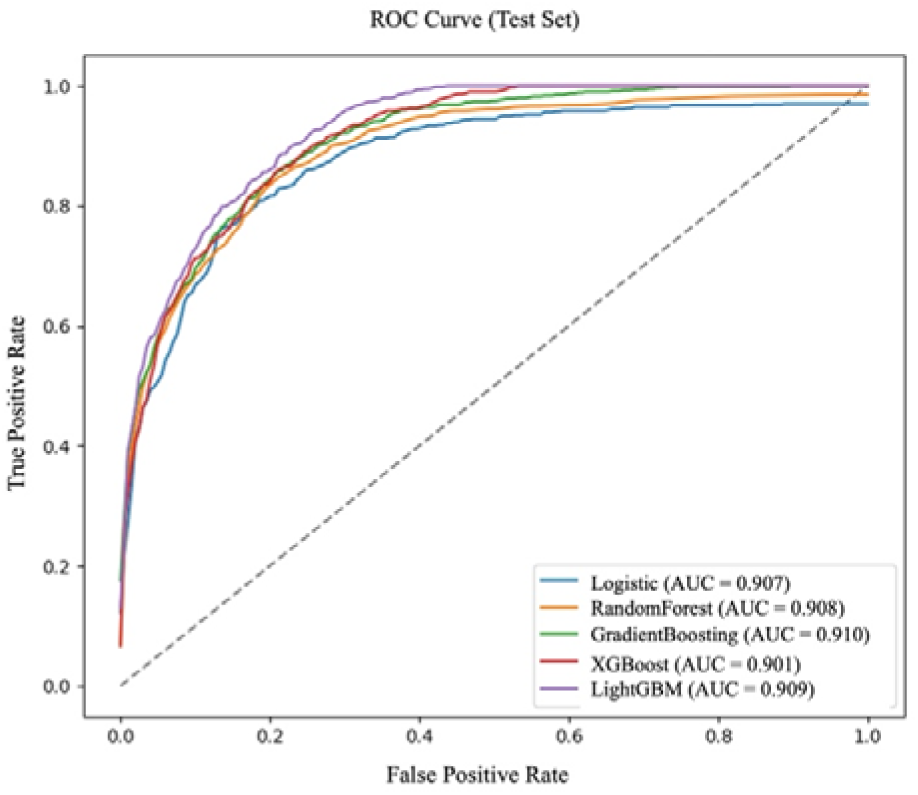
ROC Curves for Subgroup Fairness Models (Income as Sensitive Attribute) The figure presents ROC curves for models incorporating subgroup fairness analysis using income as the sensitive attribute. AUC values ranged from 0.901 to 0.910, demonstrating consistently strong predictive performance across income-based subgroups. These results indicate the models’ robustness in classifying SWB outcomes while considering potential disparities related to socioeconomic status.

**Fig. D3.**
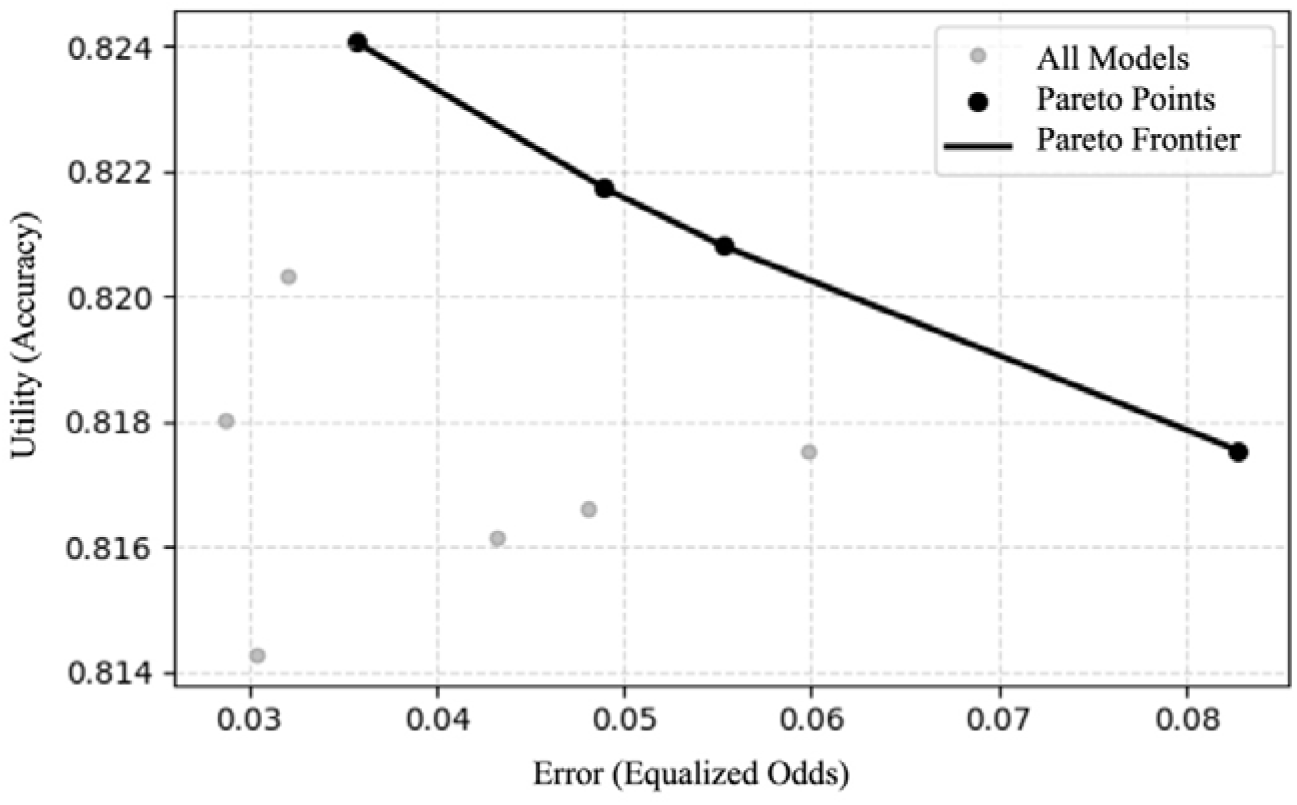
Trade-off Curve Between Fairness and Performance: A Pareto Frontier Perspective. The figure illustrates the trade-off between model utility (accuracy) and fairness (equalized odds error). Each dot represents a model, with the solid line connecting the Pareto-optimal points. The Pareto frontier highlights models that achieve the best balance between fairness and predictive performance, emphasizing the inherent tension in optimizing both objectives simultaneously.

